# Re-evaluation and Re-analysis of 152 research exomes five years after the initial report reveals clinically relevant changes in 20%

**DOI:** 10.1101/2022.10.01.22280361

**Authors:** Tobias Bartolomaeus, Julia Hentschel, Rami Abou Jamra, Bernt Popp

**Author notes:** Corresponding author **CORRESPONDING AUTHOR:** Dr. med. Bernt Popp, Institute of Human Genetics, University of Leipzig Medical Center, Leipzig, Germany, Prof. Dr. med. Rami Abou Jamra, Institute of Human Genetics, University of Leipzig Medical Center, Leipzig, Germany.

## Abstract

Iterative re-analysis of NGS results is not well investigated for published research cohorts of rare diseases.

We revisited a cohort of 152 consanguineous families with developmental disorders (NDD) reported five years ago. We re-evaluated all reported variants according to diagnostic classification guidelines or our candidate gene scoring system (AutoCaSc) and systematically scored the validity of gene-disease associations. Sequencing data was re-processed using an up-to-date pipeline for case-level re-analysis.

In 30/152 (20%) families, we identified a clinically relevant change. Thirteen previously reported (likely) pathogenic variants were re-classified as VUS/benign. In three cases, the gene-disease association (*TSEN15, NAPB*, and *FAR1*) validity was judged as limited. We identified 12 new disease causing variants. Two previously reported variants were missed by our updated pipeline due to alignment or reference issues.

Our results support the need to re-evaluate screening studies, not only the negative cases but including supposedly solved ones. This also applies in a diagnostic setting. We highlight that the complexity of computational re-analysis for old data should be weighed against the decreasing re-testing costs. Since extensive re-analysis per case is beyond the resources of most institutions, we recommend a screening procedure that would quickly identify the majority (83%) of new variants.

## INTRODUCTION

Large scale exome sequencing (ES) studies ^1^ have revealed considerable heterogeneity for sporadic neurodevelopmental disorders (NDD) with hundreds of genes affected by *de novo* variants. The NDD term is used to summarize intellectual disability (ID), developmental delay, and autism spectrum disorders (ASD). Despite the fact that current NDD cohorts with presumed recessive inheritance due to parental consanguinity are much smaller, there are more NDD genes with recessive inheritance (982 definitively associated genes with recessive inheritance vs. 527 genes with dominant inheritance, SysNDD ^2^: https://sysndd.dbmr.unibe.ch/, accessed 2022-08-25). The proportion of underlying recessive disorders in consanguineous NDD cohorts can be as high as 81%, ^3^ validating the efficacy of the traditionally chosen technique of autozygosity mapping in disease gene identification. On the other hand, *de novo*, X-linked, and compound heterozygous variants should not be overlooked in consanguineous families. ^3–5^

When compared to sporadic or dominant inheritance, recessive NDD have fewer described cases per gene, but the confidence in these associations is typically not lower. It is common to report a confirmatory case or cohort several years after the initial discovery. ^6^ Consanguineous populations are underrepresented in public databases, further exacerbating the problem. As genetic testing is often unavailable to these families, the majority of published ES studies in consanguineous families were in a research context.

In NDD cohorts examined using ES, the diagnostic yield is roughly 31-53%, ^7^ leaving 47-69% unresolved. Among the explanations are limitations of the targeted regions in ES or tertiary data analysis. ^8^ The knowledge systematized in public databases is constantly increasing, with over 300 new genetic disease associations in general ^6^ and about 160 for recessive NDD (https://sysndd.dbmr.unibe.ch/EntriesOverTime) ^2^ being identified each year. This knowledge can be automatically queried. With the recent developments in the standardization of variant interpretation, data sharing, availability of large control databases, evolved computational tools, and filtering strategies, re-assessing published NDD cohorts is intriguing because: 1) improved variant calling algorithms can demask previously concealed genomic variation; 2) diagnostic variants can be evaluated according to current standards; 3) disease associations can be confirmed or removed; and 4) previously unknown gene associations can be uncovered.

Based on these considerations and following the terminology and recommendations of the ACMG ^9^ we decided to systematically re-assess a cohort of 152 families published by some of us in 2017 using analysis steps designed to cover the above points. Our aim was to quantify the possible gains of different re-assessment levels, contrast them with their complexity, and recommend an iterative re-evaluation, re-analysis, and data-sharing scheme for NDD research screening studies. We also identify possible problems in re-analysing outdated sequencing data.

## METHODS

### Cohort and ethics statement

The analyzed cohort was previously described by Reuter and colleagues in 2017. ^10^ 152 core families with parental consanguinity and at least one child with NDD were studied. The cases were screened for genetic causes of NDD using autozygosity mapping and ES. The last data evaluation was between 2015-06-01 and 2016-08-31. Reuter et al. analyzed exome data sequentially by multiple analysts using an in-house pipeline. The Medical Faculties of Bonn and Erlangen-Nürnberg approved this cohort’s study. Some participants received compensation for travel costs.

### Re-evaluation of previously published diagnostic variants

Following the ACMG, we defined ‘re-evaluation’ as a new investigation and classification of previously reported variants. ^9^ For all variants previously reported by Reuter et al. in ‘known disease genes’, we evaluated the confidence in the gene disease association (GDA) based on the number of segregating variants, confirmatory publications, functional studies, and animal models. Next, all variants in genes with established phenotype associations were re-evaluated according to the ACMG classification ^11^ and the latest ACGS Best Practice Guidelines for Variant Classification (https://www.acgs.uk.com/media/11631/uk-practice-guidelines-for-variant-classification-v4-01-2020.pdf; Figure 1; File S2 ^12^). We submitted the results of our variant assessment to ClinVar ^13^ for all established GDAs.

**Figure 1.**
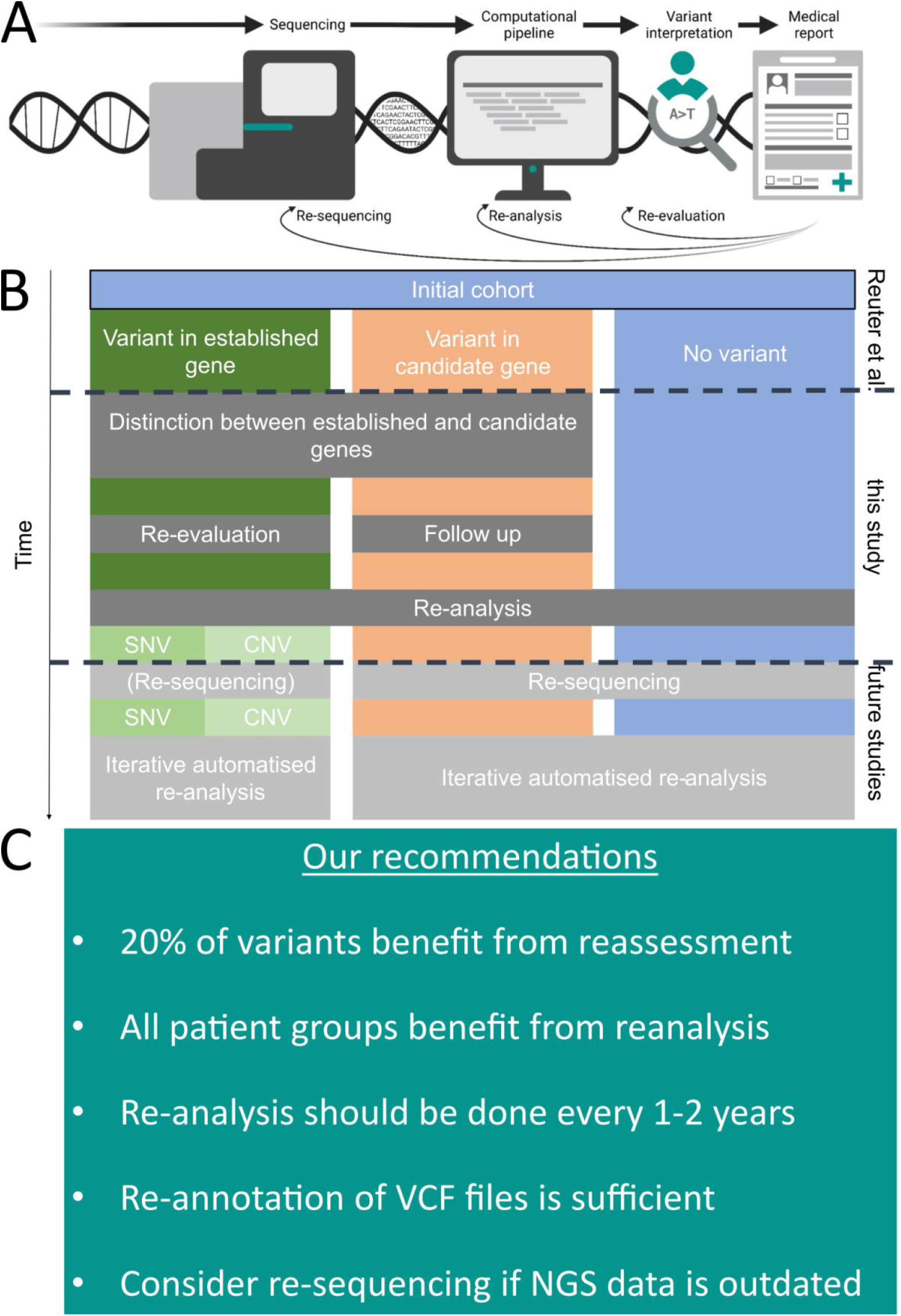
Infobox, re-evaluation and re-analysis flow diagram and recommendations. (**A**) shows an infobox summarizing the basic terms in this study (panel created with BioRender.com). (**B**) A schematic flow diagram of the previous analyses performed by Reuter et al. on the same cohort, which we re-analysed with three of the main possible outcome columns from exome screening, performed the described analyses, and finally evaluated recommendations for future pipelines incorporating automated iterative re-analysis and re-sequencing. (**C**) Infobox with our recommendations for re-evaluating and re-analysing NDD screening studies.

### Follow-up of previously published variants in candidate genes

We then assessed the association and confirmation status of all variants in genes previously reported as NDD candidates by Reuter et al. (Figure 1B; File S2 ^12^). Despite ongoing attempts by the Clinical Genome Resource (ClinGen) ^14^ to standardize GDA strengths, there is presently no agreed definition or criteria for distinguishing between established and presumed GDAs. A GDA was considered established if it was reported in at least three families and by two research groups in separate publications. Alternatively, phenotype associations were recognized if only two families or one research group reported on this gene but reliable functional data was available. Genes associated with a phenotype in the literature but not reaching this level of confidence are called ‘published candidate genes’. These genes differ from ‘candidate genes’, which are only reported in large screening cohorts. If a gene could not be reliably associated with an NDD phenotype, we used an in-house tool to score NDD candidate variants (AutoCaSc ^15^; https://autocasc.uni-leipzig.de/).

### Re-analysis of old sequencing data

In the context of our study, ‘re-analysis’ refers to the complete re-processing and evaluation of the sequencing data (Figure 1A). We curated a list of all initial study sequenced individuals and families to collect and catalog alignment files (BAM) from archive harddrives. The BAM files were converted to unaligned FASTQ files, aligned to hg38, and called using a BWA/GATK pipeline to produce a multi-sample cohort VCF (variant call format) file. We annotated the file using up-to-date tools and databases and filtered it using custom scripts to produce variant lists for manual review. We also performed coverage-based copy number (CN) calling after clustering BAM files. The resulting variant lists were manually reviewed by an experienced geneticist, visualized using the IGV browser (https://software.broadinstitute.org/software/igv/) to assess variant quality, and evaluated for biological plausibility. Following that, the variants were evaluated in diagnostic and research settings. If needed, Sanger sequencing was used for segregation analysis. Method details are described in the Supplementary notes and Tables S1 and S3.

## RESULTS

### Cohort structure

We collected sequencing data and information about age, sex, and phenotypes from 152 families (44 simplex with one, 79 multiplex with two, 24 with three, and five with four or more). The cohort characteristics are depicted in Figure 2A (details in File S2 ^12^). Most affected individuals were younger than 18 years (146/169 = 87%; one index without information). Generally, there were more affected males (male to female ratio = 107/62 ∼ 1.7; one index without information). Males accounted for 29/44 (66%) simplex family cases. Most multiplex families (44/108, 41%) had affected individuals of both sexes, while 40/108 (37%) had only males and 24/108 (22%) had only females. The affected individuals originate from 15 countries (Afghanistan, Armenia, Aserbaidschan, Egypt, Iran, Iraq, Jordan, Kazakhstan, Kuwait, Lebanon, Pakistan, Syria, Tunisia, Turkey, United Arab Emirates, Ukraine). Previously, a trio-based ES method was used to discover putative *de novo* variations in 14/44 of families with only one affected individual (File S2 ^12^; Reuter et al. stated 15/45, but we now regard MR309 as multiplex because of an affected paternal uncle).

**Figure 2.**
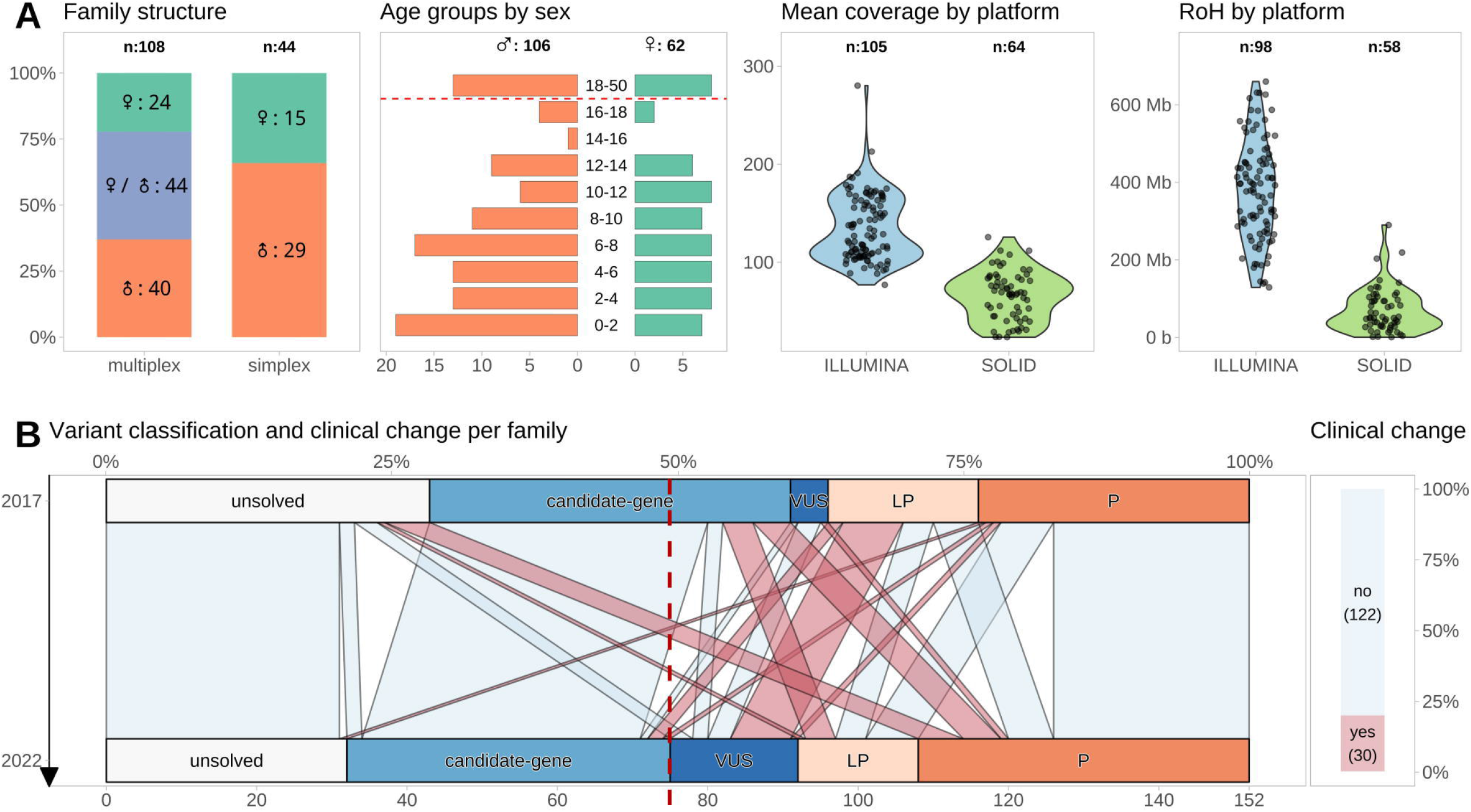
Cohort and data characteristics with results of re-evaluation. (**A**) Distribution of the 152 assessed families classified as simplex with one affected individual and multiplex with at least two affected individuals with sexdistribution of the affected individuals in these groups (first panel). The age distribution by sex in the whole cohort (note one index had no age and another had no sex data). The y-axis depicts age classes at 2-year intervals for pediatric cases, plus one class for adult index individuals. The x-axis shows the number of individuals, with females on the right (green) and males on the left (red) side (second panel). The mean coverage (third panel) and length of RoHs (fourth panel) were generally higher in samples analyzed by an Illumina platform than in samples sequenced with SOLiD technology, indicating quality differences. (**B**) The alluvial plot on the left side depicts the changes in variant classification between 2017 and 2022 as well as the results of additional identified variants. These led to clinically relevant changes in 30/152 families (20%, red; bar plot on right side colored according to the alluvial connections on the left side). The dashed line displays the shift in families where a VUS or (likely) pathogenic variant was identified. LP: likely pathogenic, P: pathogenic, RoH: run of homozygosity, VUS: variant of uncertain significance, ♀: female individual, ♂: male individual.

### Availability and quality of sequencing

We collected at least one original ES file for 169/170 (99.5%) index cases and all parental trio samples. We couldn’t retrieve 14 BAM files sequenced on a Solexa platform in March/April 2010, but for 13 of these individuals, we had subsequent ES files, so only family MR061’s index lacked original files. We added 13 new BAM files for 11 affected individuals from 10 families where ES sequencing was performed post-publication for segregation analysis. In total, we collected 216 BAM files for re-analysis, 139 from Illumina, 76 from ABI SOLiD, and one from Ion Torrent (this file was excluded because we couldn’t devise a re-processing pipeline). For samples with multiple ES attempts, we chose the best BAM, resulting in 199 files (158 original index cases, 11 new affected siblings, and 30 parental). Illumina files had significantly higher median mean read coverage (116; min: 77; max: 280) than SOLiD files (68; min: 24; max: 126) (Figure 2A and File S2 ^12^). The calculation from original hg19 to newly aligned hg38 BAMs took 17.1 CPU hours for SOLiD but only 2.9 hours for Illumina. Variant calling jobs defined by read count and covered regions took longer in Illumina BAMs (HaplotypeCaller 8.7 vs. 4.2 CPU hours). Lower coverage and quality of SOLiD files resulted in more variants remaining after filtering (median 28.5 vs. 10 homozygous calls), more CN calls (median 40.5 vs. 23.5), and more RoH calls (median 3.5 vs. 2.5); the summed RoH was shorter in the SOLiD samples (Figure 2A), likely due to heterozygous artifact calls interrupting the regions.

Two previously reported *CBS* and *FAR1* variants were missed in the recalculated data, but more sensitive variant calling allowed the identification of a lowly covered homozygous *C12orf57* variant (see discussion and Figures S3, S4 and S5).

### Novel diagnoses through re-analysis

Re-analysis revealed 12 (likely) pathogenic variants in 12 families (Table 1 and File S2 ^12^). Seven (58%) of these novel 12 diagnoses were identified in previously undiagnosed 43 families (16% diagnostic rate). This includes heterozygous *ZEB2, HNRNPH2*, a hemizygous *TAF1*, and homozygous *DEGS1, YARS1, ESPN*, and *ADD3* variants.

**Table 1.** Previously undetected variants identified in re-analysis. ^#^: the pathogenic *ESPN* variant only explains part of the phenotype (deafness); ^§^: the *ZNF292* variant does not explain the microcephaly and short stature or the NDD phenotype in the other affected individuals from this family (see dual diagnosis results in Supplementary notes); *: both affected siblings inherited the *ASXL3* variant from the unaffected father; AD: autosomal dominant, AR: autosomal recessive, dn: de novo, EEG: electroencephalogram, f: female, hemi: hemizygous, het: heterozygous, hom: homozygous, ID: intellectual disability, LP: likely pathogenic, m: male, mat: maternal, n.a.: not available, P: pathogenic, pat: paternal, VUS: variant of uncertain significance, XL: X-linked; compare File S3 ^12^

| Family | Affected | Gene | Variant | Inheritance mode | Gene classification | Variant classification | Explains phenotype | Phenotype |
| --- | --- | --- | --- | --- | --- | --- | --- | --- |
| MR152 | 2, m | *ADD3* | c.1100G>A, p.(Gly367Asp) | AR | established | P | yes | ID |
| MR333 | 1, m | *ADNP* | c.2496_2499 del, p.(Asn832Lysfs*81) | AD | established | P | yes | moderate ID, muscular hypotonia, gait disturbance, EEG abnormalities, cerebral atrophy |
| MR-DIV-01 | 2, m | *C12orf57* | c.1A>G, p.0? | AR | established | P | yes | severe ID, seizures, muscular hypotonia, short stature |
| MR-SYR-49b | 1, f | *DEGS1* | c.764A>G, p.(Asn255Ser) | AR | established | P | yes | severe ID, cerebral atrophy |
| MR128 | 1, m | *ESPN* | c.1916-1G>C, p.0? | AR | established | P | partially | severe ID, muscular hypotonia, deafness, strabismus, aplasia cutis congenita of scalp |
| MR-SYR-28 | 2, fm | *GCDH* | c.1204C>T, p.(Arg402Trp) | AR | established | P | yes | very severe ID, seizures, muscular hypotonia, limb hypertonia, spasticity, short stature, microcephaly, leukodystrophy |
| MR-TUR-05 | 1, f | *HNRNPH2* | c.616C>T, p.(Arg206Trp) | AD | established | P | yes | severe ID, myoclonus, microcephaly, muscular hypotonia, ataxia, EEG abnormalities |
| MR125 | 2, m | *YARS1* | c.1099C>T, p.(Arg367Trp) | AR | established | P | yes | ID |
| MR-SYR-49c | 1, m | *ZEB2* | c.2177_2178 del, p.(Ser726Phefs*29) | AD | established | P | yes | severe ID, seizures, microcephaly, cerebral atrophy, hypoplastic corpus callosum, atrial septal defect, pulmonic stenosis |
| MR136 | 1, f | *GRIN2A* | c.2077A>G, p.(Asn693Asp) | AD | established | LP | yes | very severe, EEG abnormalities, muscular hypotonia |
| MR073 | 3, m | TAF1 | c.2590C>T, p.(Arg864Trp) | XL | established | LP | yes | moderate ID, mental deterioration, microcephaly, nystagmus |
| MR-DIV-02 | 2, fm | ZNF292 | c.3460_3463 del, p.(Val1154Ilefs*7) | AD | established | LP | partially | mild ID, small for gestational age, short stature, microcephaly |
| MR-SYR-14 | 2, fm | ASXL3 | c.4462_4465 del, p.(Thr1488Serfs*17) | AD | established | VUS | no | mild ID, microcephaly, aggressive behavior, self-mutilation |
| MR124 | 2, m | SCN2A | c.4606A>G, p.(Ser1536Gly) | AD | established | VUS | no | very severe ID, seizures, microcephaly, short stature, cataract, cryptorchidism, pyloric stenosis, cerebral atrophy, hypoplastic corpus callosum |
| MR-SYR-06 | 5, f | *SLC35A1* | c.508-6T>C, p.? | AR | established | VUS | no | profound ID, muscular hypotonia, cerebral atrophy, seizures, short stature |
| MR145 | 2, m | *ZMIZ1* | c.418T>C, p.(Ser140Pro) | AD | established | VUS | no | moderate ID, microcephaly, short stature |
| MR-SYR-21 | 2, m | *ARHG EF6* | c.257A>C, p.(Asp86Ala) | XL | candidate | not applicable | not applicable | severe ID, limb hypertonia, microcephaly, short stature |
| MR071a | 1, f | *ZNF143* | c.44_45del, p.(Glu15Valfs*25) | AR | candidate | not applicable | not applicable | severe ID, muscular hypotonia, recurrent infections, microcephaly |

Five of the 12 novel diagnoses (42%) were found in five of the 49 families with a candidate gene variant (Figure 2B). Novel diagnoses include *ADNP, GRIN2A*, and *ZNF292* heterozygous variants and *GCDH* and *C12orf57* homozygous variants, reducing the plausibility of candidate genes (*BDH1, CLMN, FBXO11, FNDC3A, ENO2*). We likely clarified or added diagnoses in 10% of cases thought to be solved by a candidate gene.

In three families (MR124, MR-SYR-06, MR145), we found a VUS in *SCN2A, SLC35A1*, and *ZMIZ1*, respectively, but could not perform segregation analysis without parental samples. In a fourth family, MR-SYR-14, both affected siblings inherited a frameshifting variant in *ASXL3* from their unaffected father (Table 1, Figure S6). This variant is located upstream of truncating variants in *ASXL3* reported to be pathogenic, and parental mosaicism is frequent.^16^ Sanger sequencing on peripheral blood DNA indicated the variant as heterozygous, but cannot reliably discern higher-grade postzygotic events, and no tissue samples were available for the father. The healthy father’s segregation result wasn’t enough to exclude the variant, so it was classified as VUS.

### Plausibility of gene association

Reuter et al. reported 62 variants in 58 established genes in 61 families and 53 variants in 53 candidate genes in 49 families (in one family, they found a VUS and a variant in a candidate gene); 43 families had no variants. We re-evaluated all genes reported by Reuter et al. (Figure 1A). Four genes were downgraded from established to candidate status (*FAR1, TSEN15, KDM6B*, and *NAPB*; see Table 2). Four genes were previously referenced as candidate genes (*EZR, EDC3, EEF1D, NCAPD2;* see File S2 ^12^) and a disease association has been published, ^17–21^ including individuals from this cohort. However, evidence for disease association was insufficient when we re-evaluated, so we consider these eight genes published candidate genes. Six former candidate genes (*TMTC3, GALNT2, SLC44A1, TMEM94, GRM7, PTRHD1*) now have enough evidence for disease association, so we labeled them as established genes (see File S2 ^12^ for all genes and changes in gene classifications).

**Table 2.**
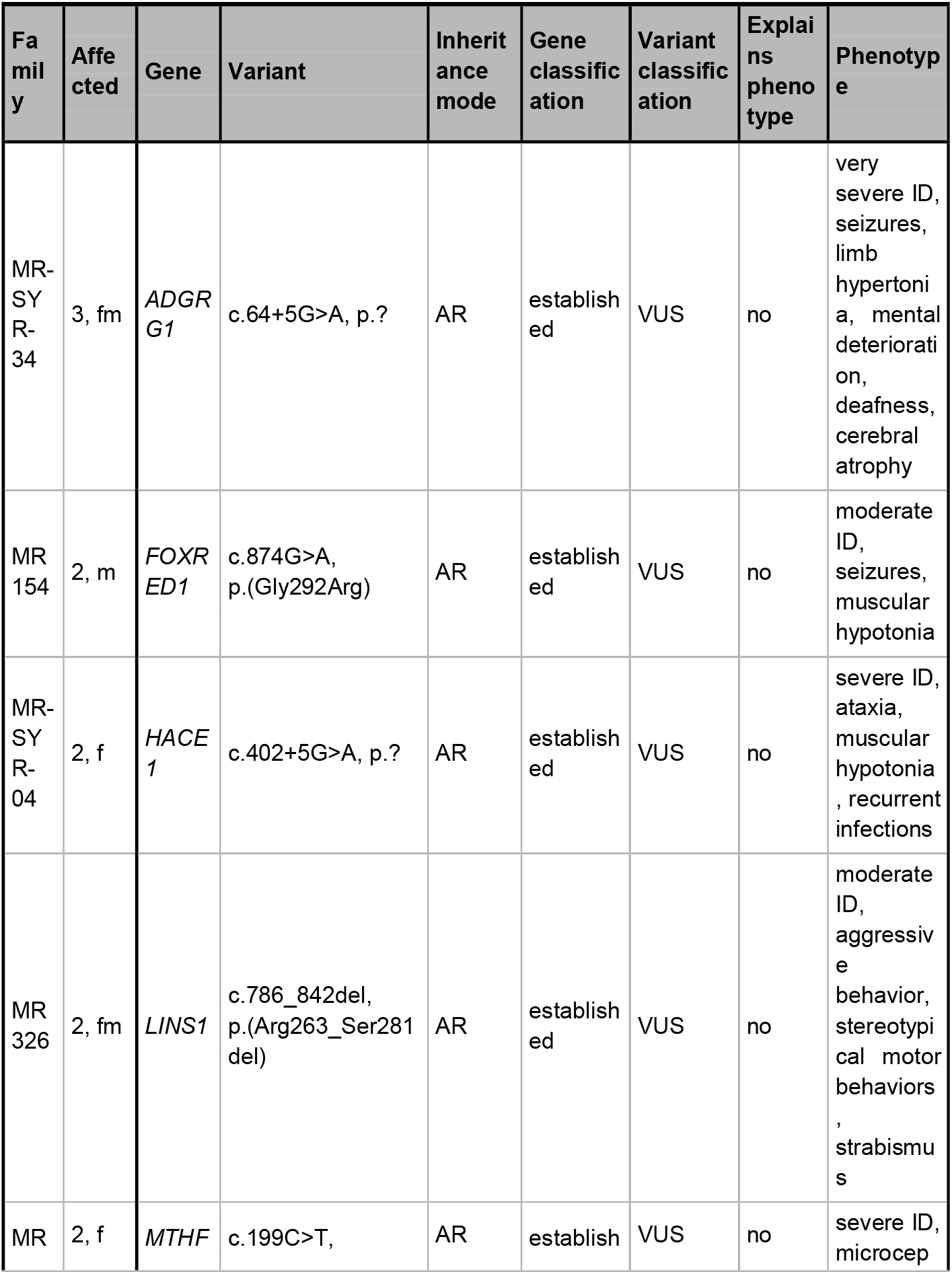
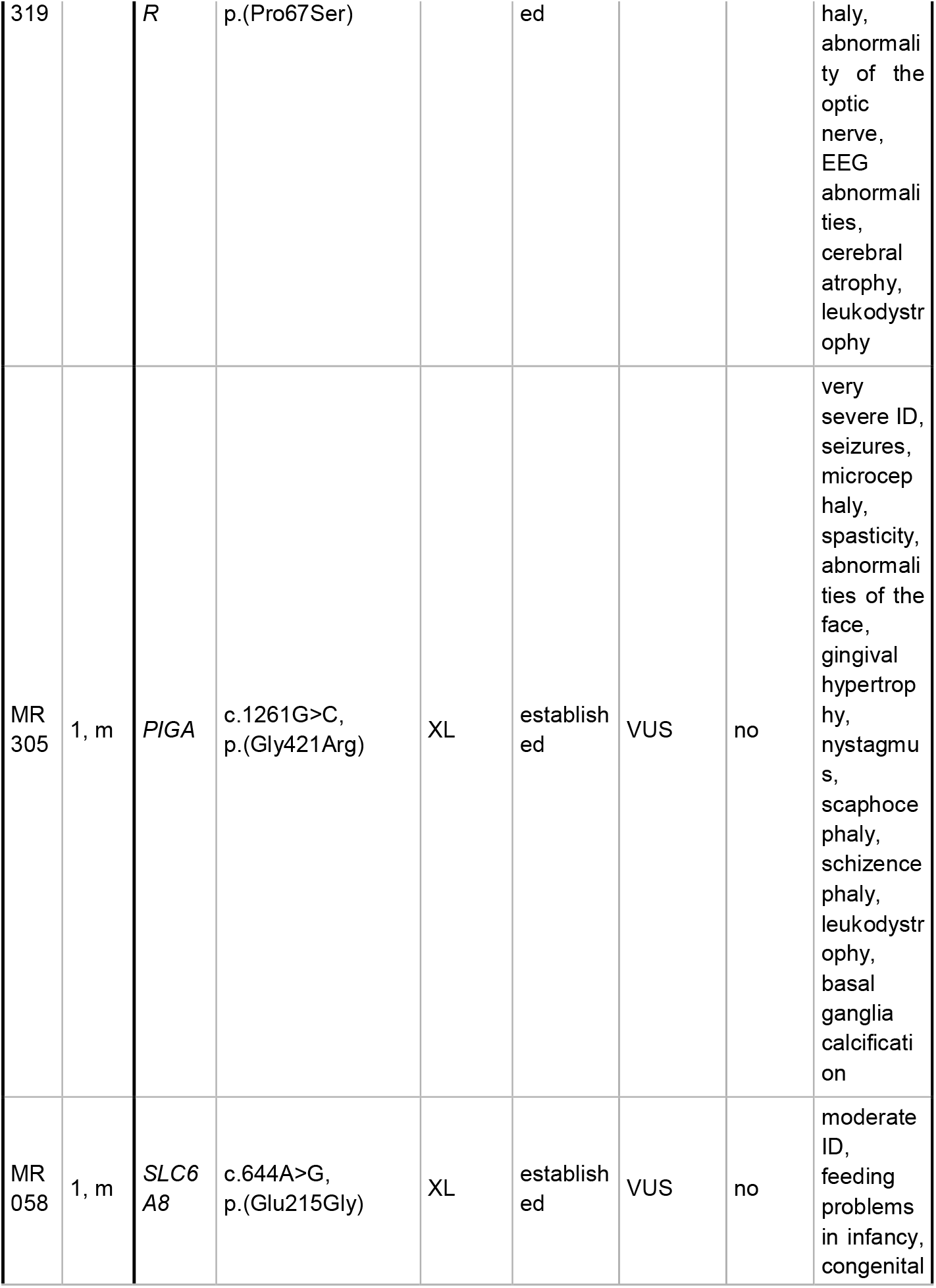
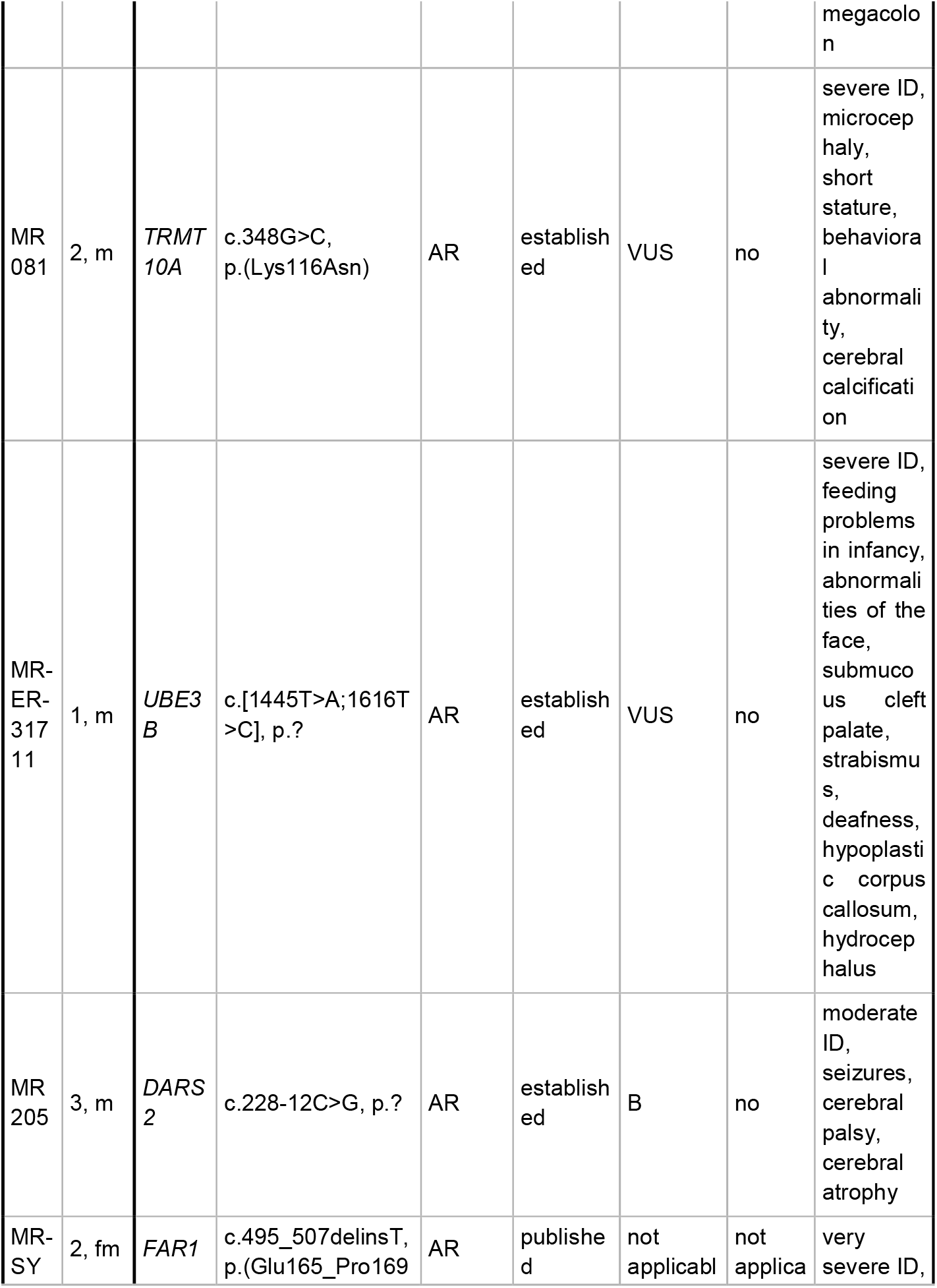
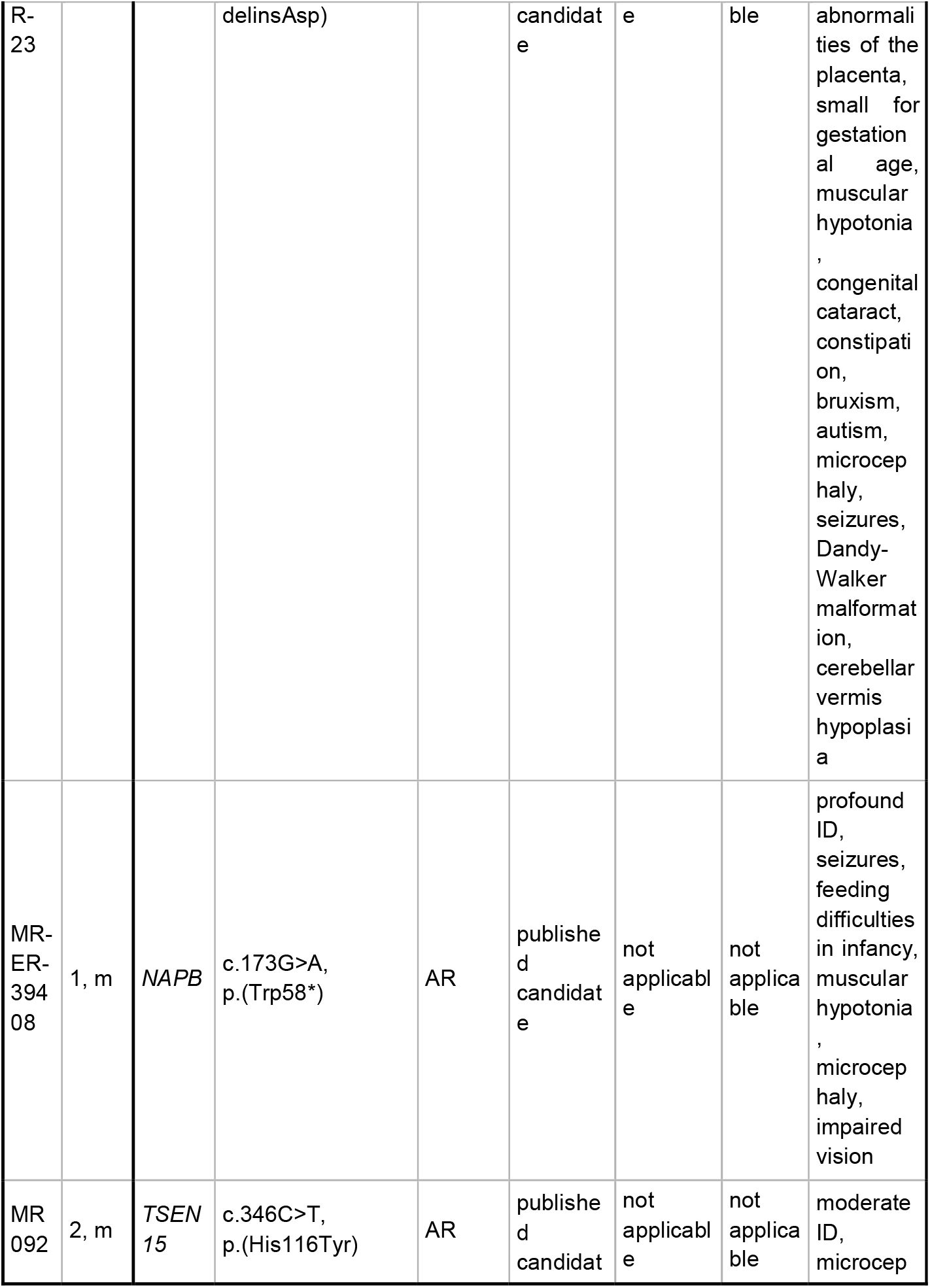
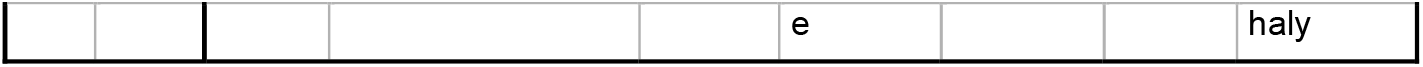
Clinical relevant changes in variant classification after re-evaluation. AR: autosomal recessive, EEG: electroencephalogram, f: female, hemi: hemizygous, hom: homozygous, ID: intellectual disability, m: male, mat: maternal, VUS: variant of uncertain significance, XL: X-linked; compare File S3 ^12^

| Fa<br>mil<br>y | Affe<br>cted | Gene | Variant | Inheri<br>tance<br>mode | Gene<br>classific<br>ation | Variant<br>classific<br>ation | Explai<br>ns pheno<br>type | Phenotyp<br>e |
| --- | --- | --- | --- | --- | --- | --- | --- | --- |
| MR-SY<br>R-34 | 3, fm | *ADGR*<br>*G1* | c.64+5G>A, p.? | AR | establish<br>ed | VUS | no | very<br>severe ID,<br>seizures,<br>limb<br>hypertoni<br>a, mental<br>deteriorati<br>on,<br>deafness,<br>cerebral<br>atrophy |
| MR<br>154 | 2, m | *FOXR*<br>*ED1* | c.874G>A,<br>p.(Gly292Arg) | AR | establish<br>ed | VUS | no | moderate<br>ID,<br>seizures,<br>muscular<br>hypotonia |
| MR-SY<br>R-04 | 2, f | *HACE*<br>*1* | c.402+5G>A, p.? | AR | establish<br>ed | VUS | no | severe ID,<br>ataxia,<br>muscular<br>hypotonia<br>, recurrent<br>infections |
| MR<br>326 | 2, fm | *LINS1* | c.786_842del,<br>p.(Arg263_Ser281<br>del) | AR | establish<br>ed | VUS | no | moderate<br>ID,<br>aggressiv<br>e<br>behavior,<br>stereotypi<br>cal motor<br>behaviors<br>,<br>strabismu<br>s |
| MR | 2, f | *MTHF* | c.199C>T, | AR | establish | VUS | no | severe ID,<br>microcep |

| Fa<br>mil<br>y | Affe<br>cted | Gene | Variant | Inherit<br>ance<br>mode | Gene<br>classific<br>ation | Variant<br>classific<br>ation | Explai<br>ns pheno<br>type | Phenotyp<br>e |
| --- | --- | --- | --- | --- | --- | --- | --- | --- |
| 319 |  | *R* | p.(Pro67Ser) |  | ed |  |  | haly,<br>abnormali<br>ty of the<br>optic<br>nerve,<br>EEG<br>abnormali<br>ties,<br>cerebral<br>atrophy,<br>leukodystro<br>phy |
| MR<br>305 | 1, m | *PIGA* | c.1261G>C,<br>p.(Gly421Arg) | XL | establish<br>ed | VUS | no | very<br>severe ID,<br>seizures,<br>microcep<br>haly,<br>spasticity,<br>abnormali<br>ties of the<br>face,<br>gingival<br>hypertrop<br>hy,<br>nystagmu<br>s,<br>scaphoce<br>phaly,<br>schizence<br>phaly,<br>leukodystro<br>phy,<br>basal<br>ganglia<br>calcificati<br>on |
| MR<br>058 | 1, m | *SLC6*<br>*A8* | c.644A>G,<br>p.(Glu215Gly) | XL | establish<br>ed | VUS | no | moderate<br>ID,<br>feeding<br>problems<br>in infancy,<br>congenital megacolo<br>n |
| MR<br>081 | 2, m | *TRMT*<br>*10A* | c.348G>C,<br>p.(Lys116Asn) | AR | establish<br>ed | VUS | no | severe ID,<br>microcep<br>haly,<br>short<br>stature,<br>behaviora<br>l<br>abnormali<br>ty,<br>cerebral<br>calcificati<br>on |
| MR-ER-317<br>11 | 1, m | *UBE3*<br>*B* | c.[1445T>A;1616T<br>>C], p.? | AR | establish<br>ed | VUS | no | severe ID,<br>feeding<br>problems<br>in infancy,<br>abnormali<br>ties of the<br>face,<br>submuc<br>ous cleft<br>palate,<br>strabismu<br>s,<br>deafness,<br>hypoplasti<br>c corpus<br>callosum,<br>hydrocep<br>halus |
| MR<br>205 | 3, m | *DARS*<br>*2* | c.228-12C>G, p.? | AR | establish<br>ed | B | no | moderate<br>ID,<br>seizures,<br>cerebral<br>palsy,<br>cerebral<br>atrophy |
| MR-SY | 2, fm | *FAR1* | c.495_507delinsT,<br>p.(Glu165_Pro169 | AR | publishe<br>d | not<br>applicabl | not<br>applica | very<br>severe ID, |

| Fa<br>mil<br>y | Affe<br>cted | Gene | Variant | Inherit<br>ance<br>mode | Gene<br>classific<br>ation | Variant<br>classific<br>ation | Explai<br>ns pheno<br>type | Phenotyp<br>e |
| --- | --- | --- | --- | --- | --- | --- | --- | --- |
| R-23 |  |  | delinsAsp) |  | candidat<br>e | e | ble | abnormali<br>ties of the<br>placenta,<br>small for<br>gestation<br>al age,<br>muscular<br>hypotonia<br>,<br>congenital<br>cataract,<br>constipati<br>on,<br>bruxism,<br>autism,<br>microcep<br>haly,<br>seizures,<br>Dandy-Walker<br>malformat<br>ion,<br>cerebellar<br>vermis<br>hypoplasi<br>a |
| MR-ER-394<br>08 | 1, m | *NAPB* | c.173G>A,<br>p.(Trp58*) | AR | publishe<br>d candidat<br>e | not applicabl<br>e | not applica<br>ble | profound<br>ID,<br>seizures,<br>feeding<br>difficulties<br>in infancy,<br>muscular<br>hypotonia<br>,<br>microcep<br>haly,<br>impaired<br>vision |
| MR<br>092 | 2, m | *TSEN*<br>*15* | c.346C>T,<br>p.(His116Tyr) | AR | publishe<br>d candidat<br>e | not applicabl<br>e | not applica<br>ble | moderate<br>ID,<br>microcep |

**Table 2 | Clinical relevant changes in variant classification after re-evaluation**
| Fa<br>mil<br>y | Affe<br>cted | Gene | Variant | Inherit<br>ance<br>mode | Gene<br>classific<br>ation | Variant<br>classific<br>ation | Explai<br>ns<br>pheno<br>type | Phenotyp<br>e |
| --- | --- | --- | --- | --- | --- | --- | --- | --- |
|  |  |  |  |  | e |  |  | haly |

### Changes in variant classification

We re-evaluated the 62 variants previously classified by Reuter et al. as (likely) pathogenic or VUS. Of the 37 pathogenic variants identified in 36 families, 34 (92%) are still considered pathogenic (27/37; 73%) or likely pathogenic (7/37; 19%), whereas one variant was changed to benign (*DARS2*), one to VUS (*LINS1*), and one was not classified because a GDA has not yet been established (*NAPB*; Table 2; Figure 2B). Of the 20 previously likely pathogenic variants, six were classified as pathogenic, four remained as likely pathogenic, eight were downgraded to VUS, and two were not classified because of an uncertain GDA (*FAR1, TSEN15*). No new evidence upgraded one of the five VUS. One was reclassified as benign (*TRAPPC9*, based on BS1, BS2, e.g., allele frequency >1% and homozygous carrier in gnomAD), and one was not classified because a recessive inheritance mode was not established (*KDM6B*; Figure 2B and File S2 ^12^). 29/62 (47%) previously reported variants weren’t in ClinVar.

### Candidate gene confirmation through matchmaking

The candidate genes identified by Reuter et al. were followed up. In 10 (19%) of the formerly 53 candidate genes an association to a neurodevelopmental disorder was meanwhile published (*GRM7, EZR, EDC3, EEF1D, TMTC3, GALNT2, SLC44A1, PTRHD1, TMEM94, NCAPD2*; File S2). For another 10 (19%) candidate genes, our upload in GeneMatcher ^22^ resulted in the first positive contact with other research groups. In four families with previous candidate genes (*BDH1, CLMN, FNDC3A, FBXO11*), a likely pathogenic variant was found, contradicting their causality for the individuals’ symptoms. For the remaining 29 (54.7%), GeneMatcher hasn’t led to contact with other researchers.

### Examples of lessons learnt

During our re-analysis, we found several potential pitfalls that we and other genetics labs can learn from. The following are three exemplary case studies.

Multiple diagnoses in one family: Family MR-SYR-49 is subdivided into three branches named ‘a’, ‘b’ and ‘c’ (Figure S1A). One variant in the candidate gene *CEP76* was reported in the ‘a’ branch. Follow up of this gene led to collaboration with other groups (ongoing research). During re-analysis, we identified a homozygous pathogenic variant in *DEGS1* (Figure S1B) in the affected girl from the ‘b’ branch. The disease association was recently published. ^18^ The affected boy from the ‘c’ branch has the autosomal dominant Mowat-Wilson syndrome due to a heterozygous pathogenic variant in *ZEB2* (OMIM #235730). To summarize, this is a large family in which three members have three unique diseases with varied inheritance patterns. The initial strategy focused on one common cause, which didn’t work.

Missing the X-chromosomal inheritance pattern: A hemizygous, likely pathogenic *TAF1* variant segregated with ID and craniofacial dysmorphism in family MR073 (Figure S1C, Table 1). We expand the mutational spectrum of this disorder by adding a *TAF1* variant that disrupts DNA interaction (Figure S1D). Due to insufficient information in population data banks and challenges in classifying inherited X-linked variants, most *TAF1* variants have unknown significance. ^24^ This family shows how consanguinity can obscure alternative inheritance patterns. We couldn’t classify this variant as pathogenic without segregation analysis, which emphasizes the need to store index, parent, and affected family DNA samples.

Identifying new variants and excluding old ones: The homozygous *TRAPPC9* variant in MR333, described previously as VUS, was reclassified as benign in this family with one affected male (criteria BS1, BS2 with >1% allele frequency and several homozygous cases in gnomAD). A *CLMN* candidate variant has also been reported. Re-analysis revealed a heterozygous (*de novo*) pathogenic *ADNP* variant that causes Helsmoortel-van der Aa syndrome (OMIM #615873). This shows that re-analysis helps more than just families with negative results. It also highlights that *de novo* variants are a possible cause, especially in singleton cases despite parental consanguinity.

## DISCUSSION

The objectives of this study were to maximize the output of our cohort of 152 consanguineous families with children who have NDD and to estimate the effort-benefit ratio so that we can make informed decisions regarding re-evaluation and re-analysis. Thus, we first re-evaluated all previously reported variants and revisited previously reported candidate genes. The raw ES data was then re-calculated, which produced a uniform callset, and then re-analysed to identify variants in known genes or novel candidate genes. For each assessment level, the added value in diagnosing new families and insights gained for variant curators can be contrasted with effort and pitfalls.

Re-evaluating 152 families resulted in clinically significant changes in 30 families (20%) due to reclassification of previously reported variants, published candidate genes, or previously undetected disease-causing variants (Figure 2B, right panel; Tables 1 and 2). While we conclude that it is generally prudent to re-evaluate all data older than five years, additional questions remain: Should this be done for all cases, or should certain levels of analysis be prioritized? Is it worthwhile to completely re-calculate old data, or should new data be generated? What timeframes are adequate for the different levels?

Thirteen (21%) of Reuter et al.’s 62 variants were reclassified clinically. The previously reported diagnostic yield for confirmed NDD genes must be retroactively adjusted from 37% to 28% (−9%). This is consistent with other groups’ findings that 40% of older variants need reinterpretation. ^25,26^ With the standardization and wider application of classification criteria, it’s less likely that this number applies to current variants. Still, it shows how important and necessary it is to re-evaluate. When variants should be re-interpreted depends on the classification used (e.g., original ACMG recommendations for Reuter et al.), but as a rule of thumb, one could recommend reinterpreting variants classified before the recent ACMG/ACGS updates in 2019/2020.

After re-analysing exome data, 12 missed (likely) pathogenic variants were found, raising the corrected diagnostic yield again from 28% to 38% (+10%). From 2010 to 2016, when the original datasets were annotated, public databases had less genetic information, such as variant frequency, reported pathogenicity, or gene-disease associations. To evaluate the many variants practically, the analysis was limited to multiple affected family members’ linkage regions. Several of the missed variants are outside of these linkage regions or are within them but could not be prioritized due to the large number of rare homozygous variants in the respective family. In addition, after Reuter et al., more gene-disease associations were published. When the majority of the data had been analyzed previously, the ‘*de novo* paradigm’ in NDD ^27^ had only recently been proposed and was not as widely accepted as it is today. The five novel *de novo* and X-linked variants found in this study went beyond the first study of this cohort and were made possible by better variant and disease databases.

Eight of 12 novel-positive families’ original annotated VCF files were available. Seven of them had the correct variant in the old data. In the eighth family (MR-DIV-01), the *C12orf57* variant was missing, likely due to poor variant quality not passing a software threshold. Considering the computational effort for the full re-analysis, a re-annotation of the VCF files with updates from gnomAD, HGMD, and ClinVar would have been sufficient to find pathogenic variants for 7/8 (88%) of these newly diagnosed families.

Although extensive re-analysis of NGS data beyond individual cases may seem impractical, we want diagnostic institutions and other research groups with access to larger screening cohorts to regularly re-analyse their published data. In contrast to our extensive re-analysis of all individual variants, a systematic re-analysis could be limited to the following steps: a) rare likely loss-of-function (LoF) variants, b) pathogenic variants in ClinVar/HGMD, and c) rare missense variants with a known pathogenic variant at the same amino acid position. This could allow computational filtering to replace time-consuming manual re-analysis. In our study, 10/12 (83%) novel pathogenic variants would have been found using this 3-step filtering method.

Compared to other recent studies of NDD in consanguineous cohorts, our 38% new diagnostic yield seems low (43.3% - 85.0%). ^25–30^ However, the wide range of diagnostic rates reflects the diversity of the cohorts studied in terms of included phenotypes and other inclusion characteristics like family structure, minimal number of affected individuals, and degree of consanguinity. Sequencing technologies and processing pipelines may also affect yield. Other groups described a share of causal CNVs between 0% - 10% ^25,27-30^, which could represent one factor of additional diagnoses. Although we called CNVs in our re-analysis, we found no new disease-causing CNVs in our cohort. This diagnostic gap is likely because the cohort was pre-analysed for CNVs before the Reuter study. Besides, the old data is probably too inhomogeneous (multiple sequencing techniques, machines, and runs throughout the project; compare Figure S2 ^12^) to have allowed detection of very small CNVs (e.g. deletion of only one exon) by coverage based methods. However, when comparing overall diagnostic yield with our results, inconsistent reporting of candidate genes is a much more important factor. In most publications, the boundary between established gene disease associations and ‘novel genes’ is not clearly defined. When strictly excluding variants in candidate genes as per our definition, the diagnostic yield in other cohorts is between 19% - 45% ^29,30^, leaving our study in the upper middle range despite its considerable heterogeneity in inclusion criteria and sequencing data quality.

In the overall view of the results of various groups ^34–39^, the re-analysis of exome data contributes to the clarification of 10 - 36% of cases. In the re-analysis of 33 consanguine families with more than two affected individuals, disease-causing variants were added in 48% (n = 16). ^8^ The described biases in the comparison with other studies of NDD in consanguineous families are further exacerbated in the comparison of the additional yield in the re-analysis of exome data. The comparison with other reports is even more confounded by our approach of re-analysing not only unsolved cases from a defined cohort, but also putatively solved cases from which we removed over-classified variants and associations, which, to our knowledge, have not been published in this way before.

Re-sequencing for this cohort was beyond the scope of our work, but the problems we encountered with re-calculation suggest possible benefits. Aligning old sequencing data to hg38 can result in ambiguous read alignment and missing variants, as with the *CBS* missense variant c.341C>T, p.(Ala114Val) (Figure S3). Some of the previous sequencing was done using SOLiD platforms. This outdated data cannot always be mapped correctly with the current mapping tools (e.g. *FAR1* in-frame indel c.495_507delinsT, compare Figure S4), leading to time consuming manual adaptation of pipelines, high computational requirements, and missed variants as well. Older research screening data is often inadequate compared to current diagnostic standards, which led to a costly validation analysis. Thus, when weighing re-analysis over re-sequencing, effort and potential problems with liftover of old target files or alignment of older read data, compatibility of new pipelines, and quality of old data should be considered. In retrospect, and assuming availability of the required funds, re-sequencing, either with diagnostic grade ES or ideally using genome sequencing, would certainly have been useful in our cohort and would have provided the additional benefit of high quality CNV analysis and detection of non-coding variants.

Our study shows the value of revisiting NDD screening data. Other groups have confirmed the need for re-analysis of sequencing data for families with no diagnostic findings and re-evaluation of reported variants. This is usually done by separating positive families from those with reported candidate genes and negative families. This leads to path dependency, where presumed positive families are considered to be conclusive. We demonstrate here that all groups benefit from re-analysis and that considerations of diagnostic yield should not be separated from re-evaluation of the entire cohort.

Based on our experience, the most promising and practical step is re-analysing existing variant calls by annotating and filtering them with the latest information and re-evaluating such identified variants together with previously reported ones according to current standards and new literature. A sensible timeframe to perform this iterative step in research cohorts could be about one to two years. Stringent submission and updates of all reported variants in known genes into public databases like ClinVar can’t be stressed enough in this regard. In contrast, computationally expensive re-analysis steps, like new alignments and variant calling, seem to provide no justifiable gain and should be reserved for re-calculations to normalize and integrate a sequenced cohort into large public datasets. While re-sequencing was not directly investigated in our study, several markers suggest a technological ‘tipping-point’ (e.g. current exome enrichment kits sequenced with 150 bp paired-end reads) has been surpassed for some of the sequencing data in this NDD cohort. The next tipping points for genetics screening could be genome sequencing, long-read sequencing, and functional sequencing like RNA-seq and Methyl-seq.

Our results support the need to regularly re-evaluate all NDD screening cases using re-annotation and efficient filtering. Rare disease studies should also incorporate biobanking protocols to enable novel re-sequencing steps.

## Data Availability

All data generated or analyzed can be found either at the publisher's website or has been uploaded to Zenodo (File S2, S3: DOI: 10.5281/zenodo.6206868).

https://zenodo.org/record/7113408

## WEB RESOURCES

AutoCaSc: https://autocasc.uni-leipzig.de/

ClinVar: https://www.ncbi.nlm.nih.gov/clinvar/

gnomAD browser: http://gnomad.broadinstitute.org/

SysNDD database: https://sysndd.dbmr.unibe.ch/

GeneMatcher: https://genematcher.org/

trRosetta: https://yanglab.nankai.edu.cn/trRosetta

## ACKNOWLEDGEMENTS

We thank all the involved families for participating in this study. B.P. is supported by the Deutsche Forschungsgemeinschaft (DFG) through grant PO2366/2–1.

## AVAILABILITY OF DATA AND MATERIALS

All data generated or analyzed can be found either at the publisher’s website or has been uploaded to Zenodo (File S2, S3: DOI: 10.5281/zenodo.7113408).

## SUPPORTING INFORMATION

File S1: supplementary methods, figures and tables

## AUTHORS’ CONTRIBUTIONS

B.P., T.B. and R.A.J. conceived the initial study concept. B.P. and R.A.J. collected and cataloged the sequencing files prior to re-analysis. T.B. and B.P. analyzed the variant data, created Figures and all supplementary materials. J.H. performed segregation analyses. T.B., R.A.J. and B.P. wrote and edited the manuscript. All authors reviewed and commented on the final draft manuscript.

## REFERENCES

1. Deciphering Developmental Disorders Study. Prevalence and architecture of de novo mutations in developmental disorders. Nature. 2017;542(7642):433–438. doi:10.1038/nature21062

2. Kochinke K, Zweier C, Nijhof B, et al. Systematic Phenomics Analysis Deconvolutes Genes Mutated in Intellectual Disability into Biologically Coherent Modules. The American Journal of Human Genetics. 2016;98(1):149–164. doi:10.1016/j.ajhg.2015.11.024

3. Anazi S, Maddirevula S, Faqeih E, et al. Clinical genomics expands the morbid genome of intellectual disability and offers a high diagnostic yield. Mol Psychiatry. 2017;22(4):615–624. doi:10.1038/mp.2016.113

4. Jalkh N, Corbani S, Haidar Z, et al. The added value of WES reanalysis in the field of genetic diagnosis: lessons learned from 200 exomes in the Lebanese population. BMC Med Genomics. 2019;12(1):11. doi:10.1186/s12920-019-0474-y

5. Al-Nabhani M, Al-Rashdi S, Al-Murshedi F, et al. Reanalysis of exome sequencing data of intellectual disability samples: Yields and benefits. Clin Genet. 2018;94(6):495–501. doi:10.1111/cge.13438

6. Boycott KM, Rath A, Chong JX, et al. International Cooperation to Enable the Diagnosis of All Rare Genetic Diseases. The American Journal of Human Genetics. 2017;100(5):695–705. doi:10.1016/j.ajhg.2017.04.003

7. Srivastava S, Love-Nichols JA, Dies KA, et al. Meta-analysis and multidisciplinary consensus statement: exome sequencing is a first-tier clinical diagnostic test for individuals with neurodevelopmental disorders. Genet Med. 2019;21(11):2413–2421. doi:10.1038/s41436-019-0554-6

8. Shamseldin HE, Maddirevula S, Faqeih E, et al. Increasing the sensitivity of clinical exome sequencing through improved filtration strategy. Genet Med. 2017;19(5):593–598. doi:10.1038/gim.2016.155

9. Deignan JL, Chung WK, Kearney HM, Monaghan KG, Rehder CW, Chao EC. Points to consider in the reevaluation and reanalysis of genomic test results: a statement of the American College of Medical Genetics and Genomics (ACMG). Genetics in Medicine. 2019;21(6):1267–1270. doi:10.1038/s41436-019-0478-1

10. Reuter MS, Tawamie H, Buchert R, et al. Diagnostic Yield and Novel Candidate Genes by Exome Sequencing in 152 Consanguineous Families With Neurodevelopmental Disorders. JAMA Psychiatry. 2017;74(3):293. doi:10.1001/jamapsychiatry.2016.3798

11. Richards S, Aziz N, Bale S, et al. Standards and guidelines for the interpretation of sequence variants: a joint consensus recommendation of the American College of Medical Genetics and Genomics and the Association for Molecular Pathology. Genet Med. 2015;17(5):405–424. doi:10.1038/gim.2015.30

12. Popp, Bernt, Bartolomaeus, Tobias. Data files for manuscript “Re-evaluation and Re-analysis of 152 research exomes five years after the initial report reveals clinically relevant changes in 20%.” Published online September 23, 2022. doi:10.5281/zenodo.7113408

13. Landrum MJ, Lee JM, Benson M, et al. ClinVar: improving access to variant interpretations and supporting evidence. Nucleic Acids Res. 2018;46(D1):D1062–D1067. doi:10.1093/nar/gkx1153

14. Strande NT, Riggs ER, Buchanan AH, et al. Evaluating the Clinical Validity of Gene-Disease Associations: An Evidence-Based Framework Developed by the Clinical Genome Resource. Am J Hum Genet. 2017;100(6):895–906. doi:10.1016/j.ajhg.2017.04.015

15. Lieberwirth JK, Büttner B, Klöckner C, Platzer K, Popp B, Abou Jamra R. AutoCaSc: Prioritizing candidate genes for neurodevelopmental disorders. Human Mutation. Published online September 14, 2022:humu.24451. doi:10.1002/humu.24451

16. Schirwani S, Hauser N, Platt A, et al. Mosaicism in ASXL3-related syndrome: Description of five patients from three families. Eur J Med Genet. 2020;63(6):103925. doi:10.1016/j.ejmg.2020.103925

17. Marafi D, Mitani T, Isikay S, et al. Biallelic GRM7 variants cause epilepsy, microcephaly, and cerebral atrophy. Ann Clin Transl Neurol. 2020;7(5):610–627. doi:10.1002/acn3.51003

18. Riecken LB, Tawamie H, Dornblut C, et al. Inhibition of RAS activation due to a homozygous ezrin variant in patients with profound intellectual disability. Hum Mutat. 2015;36(2):270–278. doi:10.1002/humu.22737

19. Martin CA, Murray JE, Carroll P, et al. Mutations in genes encoding condensin complex proteins cause microcephaly through decatenation failure at mitosis. Genes Dev. 2016;30(19):2158–2172. doi:10.1101/gad.286351.116

20. Ugur Iseri SA, Yucesan E, Tuncer FN, et al. Biallelic loss of EEF1D function links heat shock response pathway to autosomal recessive intellectual disability. J Hum Genet. 2019;64(5):421–426. doi:10.1038/s10038-019-0570-z

21. Khodadadi H, Azcona LJ, Aghamollaii V, et al. PTRHD1 (C2orf79) mutations lead to autosomal-recessive intellectual disability and parkinsonism. Mov Disord. 2017;32(2):287–291. doi:10.1002/mds.26824

22. Sobreira N, Schiettecatte F, Valle D, Hamosh A. GeneMatcher: a matching tool for connecting investigators with an interest in the same gene. Hum Mutat. 2015;36(10):928–930. doi:10.1002/humu.22844

23. Pant DC, Dorboz I, Schluter A, et al. Loss of the sphingolipid desaturase DEGS1 causes hypomyelinating leukodystrophy. J Clin Invest. 2019;129(3):1240–1256. doi:10.1172/JCI123959

24. Martin HC, Gardner EJ, Samocha KE, et al. The contribution of X-linked coding variation to severe developmental disorders. Nat Commun. 2021;12(1):627. doi:10.1038/s41467-020-20852-3

25. Xiang J, Yang J, Chen L, et al. Reinterpretation of common pathogenic variants in ClinVar revealed a high proportion of downgrades. Sci Rep. 2020;10(1):331. doi:10.1038/s41598-019-57335-5

26. SoRelle JA, Thodeson DM, Arnold S, Gotway G, Park JY. Clinical Utility of Reinterpreting Previously Reported Genomic Epilepsy Test Results for Pediatric Patients. JAMA Pediatr. 2019;173(1):e182302. doi:10.1001/jamapediatrics.2018.2302

27. Huang K. De novo paradigm: the ultimate answer to the paradox in mental retardation? Clin Genet. 2011;79(5):427–428. doi:10.1111/j.1399-0004.2011.01630.x

28. Hu H, Kahrizi K, Musante L, et al. Genetics of intellectual disability in consanguineous families. Mol Psychiatry. 2019;24(7):1027–1039. doi:10.1038/s41380-017-0012-2

29. Alazami AM, Patel N, Shamseldin HE, et al. Accelerating novel candidate gene discovery in neurogenetic disorders via whole-exome sequencing of prescreened multiplex consanguineous families. Cell Rep. 2015;10(2):148–161. doi:10.1016/j.celrep.2014.12.015

30. Karaca E, Harel T, Pehlivan D, et al. Genes that affect brain structure and function identified by rare variant analyses of Mendelian neurologic disease. Neuron. 2015;88(3):499–513. doi:10.1016/j.neuron.2015.09.048

31. Monies D, Abouelhoda M, Assoum M, et al. Lessons Learned from Large-Scale, First-Tier Clinical Exome Sequencing in a Highly Consanguineous Population. Am J Hum Genet. 2019;105(4):879. doi:10.1016/j.ajhg.2019.09.019

32. AluDewik N, Mohd H, AluMureikhi M, et al. Clinical exome sequencing in 509 Middle Eastern families with suspected Mendelian diseases: The Qatari experience. Am J Med Genet A. 2019;179(6):927–935. doi:10.1002/ajmg.a.61126

33. Eaton A, Hartley T, Kernohan K, et al. When to think outside the autozygome: Best practices for exome sequencing in “consanguineous” families. Clinical Genetics. 2020;97(6):835–843. doi:10.1111/cge.13736

34. Schmitz-Abe K, Li Q, Rosen SM, et al. Unique bioinformatic approach and comprehensive reanalysis improve diagnostic yield of clinical exomes. Eur J Hum Genet. 2019;27(9):1398–1405. doi:10.1038/s41431-019-0401-x

35. Li J, Gao K, Yan H, et al. Reanalysis of whole exome sequencing data in patients with epilepsy and intellectual disability/mental retardation. Gene. 2019;700:168–175. doi:10.1016/j.gene.2019.03.037

36. Baker SW, Murrell JR, Nesbitt AI, et al. Automated Clinical Exome Reanalysis Reveals Novel Diagnoses. The Journal of Molecular Diagnostics. 2019;21(1):38–48. doi:10.1016/j.jmoldx.2018.07.008

37. Wenger AM, Guturu H, Bernstein JA, Bejerano G. Systematic reanalysis of clinical exome data yields additional diagnoses: implications for providers. Genetics in Medicine. 2017;19(2):209–214. doi:10.1038/gim.2016.88

38. Eldomery MK, Coban-Akdemir Z, Harel T, et al. Lessons learned from additional research analyses of unsolved clinical exome cases. Genome Med. 2017;9(1):26. doi:10.1186/s13073-017-0412-6

39. Wright CF, McRae JF, Clayton S, et al. Making new genetic diagnoses with old data: iterative reanalysis and reporting from genome-wide data in 1,133 families with developmental disorders. Genetics in Medicine. 2018;20(10):1216–1223. doi:10.1038/gim.2017.246

