## Supplementary notes for "Re-evaluation and Re-analysis of 152 research exomes five years after the initial report reveals clinically relevant changes in 20%"

[*] Corresponding author

### SUPPLEMENTARY METHODS

#### New alignment from old BAM files

For all alignments, we used the 1000 genomes GRCh38 reference with decoy and HLA contigs downloaded from the NCBI FTP (File Transfer Protocol) server. The previously aligned BAM (Binary SAM (Sequence Alignment Map)) files were used as input for the new alignment pipeline divided by the respective sequencing platform.

For the ABI (Applied Biosystems, Waltham, Massachusetts, USA) SOLiD color-space (reads contain information on consecutive dinucleotides which are measured and represented using four possible colors) BAMs, previously aligned to hg19 with the proprietary ABI LifeScope software pipeline, we extracted the color-space read information from the CS (original color-space sequence) and CQ (original color-space quality values) tags using the samtools (version 1.10) 'sort' and 'fastq' commands with custom Bash and AWK scripts to generate reverted color-space FASTQ (extension of the FASTA format incorporating base qualities) files. These FASTQ files were split every 1,000,000 reads for subsequent alignment parallelization. We used the free version of novoalignCS (V1.07.00; Novocraft Technologies, Petaling Jaya, Malaysia) with standard settings and parallelized it on the split FASTQ input files using GNU Parallel [1]. The resulting output was sorted, merged, and indexed with samtools [2] to generate the new per-sample BAM files.

For the Illumina (San Diego, CA, USA) base-space (reads contain information on consecutive nucleotides) BAMs, previously aligned to hg19 with different versions of GATK/BWA based pipelines, we designed a workflow based on current GATK [3] recommendations [4] for new alignment of BAM files. First, we used the Picard Tools (version 2.21.2-0) 'RevertSam' command to generate an unmapped BAM file. Adapters were marked using Picard Tools 'MarkIlluminaAdapters'. The resulting unmapped BAM files with marked adapters were then converted to interleaved FASTQ files using Picard 'SamToFastq'. These were used as inputs for BWA-MEM [5] (version 0.7.17-r1188). The BWA-MEM SAM output was merged with the data in the respective unmapped BAM file using 'MergeBamAlignment' from Picard Tools to generate the new per-sample BAM files.

Duplicates in the merged SOLiD and Illumina BAM files were then marked with Picard Tools 'MarkDuplicatesWithMateCigar' to produce the final analysis ready hg38/GRCh38 BAM files (for alignment statistics see File S2 sheet 'BAMs').

#### A combined target file for all exome designs used

A merged BED (Browser Extensible Data) file was generated by first merging the three official BED hg19 target files (downloaded from Agilent Earray on 2019-12-23; Agilent Technologies, Santa Clara, California, United States), then lifting this file to hg38 using CrossMap.py [6] (version v0.2.7) and finally padding with 100 bp on each interval side.

#### Quality control of exome data

Quality control on the resulting analysis ready hg38 BAM files was performed using FastQC (version v0.11.9) and qualimap [7] (version v.2.2.2) and the above described merged BED files but without padding (see File S2 sheet 'BAMs' for coverage statistics).

**SNV/indel calling and annotation**

The final merged and deduped BAM files were called individually using GATK (version 4.1.4.1) HaplotypeCaller [8] to produce per sample genomic VCFs (GVCFs). Subsequently, we used gatk 'GenomicsDBImport' parallelized on the hg38 scattered calling intervals downloaded from the broad resources bundle to merge the GVCFs. Joint genotyping was done with gatk 'GenotypeGVCFs' on the same calling intervals. The resulting per interval cohort VCFs were merged into one VCF using Picard's 'MergeVcfs'. In the following steps, this cohort VCF was subset to the combined target BED file. The target VCF was subset into two files containing SNPs and other variants (indels, MNVs), respectively. These were used to recalibrate the variant quality scores using the recommended setting of gatk 'VariantRecalibrator' and apply the models using gatk 'ApplyVQSR'. The resulting two VCFs were merged again using the Picard command 'MergeVcfs'. Indel variants were left normalized and multiallelic variants were split using the gatk 'LeftAlignAndTrimVariants'. The final cohort VCF was compressed with gzip [9] and then indexed using Tabix [10].

**VCF annotation**

For annotation of the cohort VCF, we used the SnpEff [11] and SnpSift [12] (version 4.3t) annotation software packages. After genetic and functional effects were annotated with SnpEff, we next annotated various databases containing computational predictions, variant frequencies in control populations, and curated variant pathogenicity with SnpSift. These datasets included dbNSFP [13] (version 4.0a), SPIDEX [14] (version 1.0), dbscSNV [15] (version 1.1), CADD [16] PHRED scores (version v1.5), gnomAD [17] variant frequencies (version exomes r2.1.1, version genomes r3.0) and TOPMED/BRAVO [18] (Freeze5) variant frequencies. Additionally, the clinical significance from ClinVar [19] (version 2020-01-27) and variant type information from HGMD (Human Gene Mutation Database; version 2019.3; QIAGEN, Venlo, The Netherlands) were annotated. All additional annotation files were downloaded from the respective websites and prepared as SnpSift compatible input using custom Bash and AWK scripts. Database files only available as hg19/ GRCh37 versions (SPIDEX, dbscSNV, HGMD) were lifted to hg38 using CrossMap.py. For faster annotation, the large CADD, gnomAD and BRAVO files were subset to the padded BED file previously used to subset the cohort VCF file.

After variant level annotation, we additionally annotated gene level information from dbNSFP and GEVIR [20] gene ranking information (downloaded on 2020-02-03 from http://www.gevirank.org/static/files/gene_ranking.csv) using a custom R script.

**SN/indel variant filtering and review**

The annotated cohort VCF file was filtered for recessive (homozygous and compound-heterozygous) and dominant or X-chromosomal variation with a set of Bash scripts using the SnpSift 'filter' command and parallelized using GNU Parallel.

For homozygous recessive inheritance, we filtered the cohort VCF file for variants called homozygous with >= 3 reads, an AF (allele frequency) <0.02 in the cohort, anAF <0.01 in gnomAD/BRAVO with no homozygous occurrences allowed and QUAL (representing the PHRED scaled probability that the respective site has no variant) >=250 or a PASS filter (e.g.

a flag in the VCF FILTER column indicating that the respective variant passed all filters applied like in the variant recalibration step). Variants reported as benign or likely benign (without contradiction) in ClinVar were excluded. All variants reported as (likely) pathogenic in ClinVar or HGMD were kept. Likely gene disrupting variants were included if they were annotated as HIGH (SnpEff effect impact category comprising mostly protein truncating variants) or had a LOF (loss-of-function) or NMD (nonsense mediated RNA-decay) probability of > 0.5 or were annotated as 'stop', 'inframe' or 'initiator' variant. Missense variants were kept if they had a CADD PHRED >=15. Splice variants not affecting the canonical splice motives (+/-2 bp from the intron junction) were included if they had CADD PHRED >=15 and one of the three splice scores (SPIDEX, dbscSNV-ada, and -rf) passed the respective threshold (spidex_dpsi_zscore <-2.0, dbscSNV_ada_score >0.6, dbscSNV_rf_score >0.6).

Filtering for compound heterozygous recessive, dominant, or X-chromosomal inheritance was performed analogously, but heterozygous variants required >=4 reads. For the dominant and X-chromosomal inheritance patterns, we also decreased the AF in the cohort to <0.01 and in public databases to <0.001. Additionally, we subset these analyses to only known or candidate genes from the SysID [21] database (downloaded from the official website on 2020-01-11) to further reduce the large number of variants requiring review.

Due to the low sequencing quality of SOLiD exomes, only variants that 1) are reported as pathogenic in HGMD or ClinVar, 2) have putative high impact according to their Sequence Ontology term (SnpEff), or 3) are missense with read coverage >19 reads, have a MAF (minor allele frequency) in gnomAD consistent with the disease, and an alternative allele fraction consistent with the mode of inheritance (>0.3 in AD phenotypes, >0.8 in AR phenotypes). In other exome data from Illumina platforms, all variants were assessed except heterozygous variants with a MAF in gnomAD >0.01 or with ambiguous (<0.2 or >0.8) alternative allele fraction (AF). All variants reported herein were manually checked in Integrative Genomics Viewer (IGV) [22] regarding their quality and resequenced with Sanger if equivocal. The available clinical information of the individuals was compared with associated phenotypes in OMIM (Online Mendelian Inheritance in Man) [23] or in the HGMD, or with literature searches in PubMed for associated diseases. Associated phenotypes in OMIM, HGMD, and Pubmed were reviewed regarding a possible phenotypic overlap and matching pathomechanism. Classification was done according to the ACMG system and the latest updates published by the ClinGen consortium.

**CN variant calling**

Because multiple sequencing technologies and exome enrichment kits have been used throughout the different sequencing runs in this project, we first performed a PCA (principal component analysis) and k-means clustering analysis in R (packages 'tidyverse', 'broom', 'scales') on the coverage statistics computed through the 'multibamqc' command of qualimap. Based on the clustering results (Figure S1) we generated one-vs-all work lists for all the samples in each respective cluster.

For each sample, read coverage based copy number (CN) variant calling from exome data was then performed on the hg38 BAM-files utilizing the CNVkit [24] (version 0.9.6.dev0) 'batch' command. The remaining samples in the respective cluster were used as controls for each sample. Except for the 'drop-low-coverage' flag, standard settings were used in CNVkit. For the SOLiD BAMs, we used the 'amplicon' mode because off-target reads had been previously removed in the LifeScope pipeline and were thus not available for antitarget coverage computation. For the Illumina BAMs, we used the 'hybrid' mode with anti-target coverage computation. The CNVkit 'call' command was used with recommended threshold settings (-2.0, -0.41, 0.32, 0.80, 1.1) to call CN segments. Results were visualized with the 'scatter' and 'heatmap' functions in CNVkit. A custom script (grep, sed, AWK) was used to generate an analysis table containing the called CN segments and this was then annotated with information from OMIM and SysID using a custom R script (see File S3 sheet 'CNVkit_one-vs-all').

**CN variant filtering and review**

We manually evaluated all CN calls aggregated into an Excel table from the above pipeline, following a set of criteria defined after inspecting the distribution of quality measurements and the number of calls per sample. We therefore manually reviewed all CN variants with a homozygous call (CN 0), all variants with log2 values <-0.8 or >0.7, all variants falling into known microdeletion regions from DECIPHER [25], and known OMIM disease genes with a NDD association (SysID). CN variants meeting these criteria were manually evaluated using the IGV browser to visually compare coverage profiles and allele distributions with matched BAM files from similar sequencing technologies. None of the likely real CN variants from this analysis in the cohort were evaluated as relevant for the phenotype of the respective individual. For details, see File S3 sheet 'CNVkit_one-vs-all'.

**Calling runs of homozygosity from exome data**

We applied the BCFtools [26] 'roh' command to call run of homozygosity (RoH) from the annotated cohort VCF files. Using a custom R script, the results were annotated with sample identifiers, allowing comparison of sequencing technologies. For details, see File S3 sheet 'HomozygosityMappingRegions_hg38'.

**Sanger sequencing for segregation analysis and variant validation**

All newly identified variants were confirmed by targeted PCR and Sanger sequencing on the original DNA samples. Primers (see Table S1 for sequences) were designed with the ExonPrimer implementation of Primer3 [27(p3)]. Segregation was performed on all available DNAs from the respective family with the same primers. PCR products were visualized by gel electrophoresis. Sanger-sequence traces were analyzed using UGENE [28] (version 36; Unipro, Novosibirsk, Russia).

**Genetic fingerprinting**

Sample identity was confirmed for all potential *de novo* variants using genetic fingerprinting methods. We either used the PowerPlexTM21 multiplex PCR assay (20 polymorphic markers; Promega, Fitchburg, USA) analyzed on an automatic capillary sequencer (ABI3500dx; Thermo Fisher Scientific, Waltham, USA) or the EasySeq Human ID and Sample Tracking kit (36 SNPs; Nimagen, Nijmegen, Netherlands) sequenced on an NextSeq 550 System (Illumina, San Diego, California, United States).

### Analysis of selected missense variants in their 3D protein structure

To analyze the exemplary and instructive newly identified missense variants in TAF1 and DEGS1 in the tertiary protein structure, we first searched the protein data bank (RCSB PDB) for crystal structures. For *TAF1*, this search identified the EM-structure 5FUR [29]. As our search identified no reliable PDB model covering the DEGS1 variant, we applied the trRosetta [30] tool to perform a *de novo* protein structure prediction using the protein sequence in FASTA format. Both structures were visualized with the Pymol molecular visualization software (Open Source version 2.5.0; Schrödinger LLC, New York, USA).

### Data analysis and plotting

We analyzed and graphically processed all the tabular data compiled in Excel (Microsoft Corporation, Redmond, Washington, USA) using the R language (Version 4.1.3) and RStudio (Version 2022.02.3). We used the libraries 'cowplot', 'fs', 'fuzzyjoin', 'ggalluvial', 'jsonlite', 'RColorBrewer', 'readxl', 'RMySQL', 'RSQLite', 'showtext' and 'tidyverse'.

**SUPPLEMENTARY RESULTS**

### Dual diagnoses

A dual diagnosis was previously reported in one family (MR100). In two additional families, we assume a dual diagnosis since the pathogenic variants detected only explain fractions of the individual's symptoms. In one individual from MR-DIV-02 a likely pathogenic variant in *ZNF292* was later detected (published as 19-012 [31]). Besides mild ID, she also presented with short stature and microcephaly, while other individuals with pathogenic *ZNF292* variants did not. Together with the affected sibling that does not carry this *de novo ZNF292* variant, we expect an additional variant might cause short stature and microcephaly in the family. Reuter et al. found a homozygous missense variant in NCAPD2 (File S2) in this individual. Splice variants predicted to truncate *NCAPD2* were recently reported to cause microcephaly, mild ID, and reduced height. [32,33] In family MR128, the affected member has severe ID, muscular hypotonia, deafness, strabismus, and aplasia cutis congenita of the scalp. The identified pathogenic variant in ESPN only explains the deafness but is not associated with NDD or the other phenotypes observed in the family, leading to the assumption that other genetic causes remain undetected despite relatively high ES quality in this family, which indicates that genome sequencing might be a valuable next step.

### SUPPLEMENTARY FIGURES

#### Figure S1 | Selected clinical examples for previously missed variants

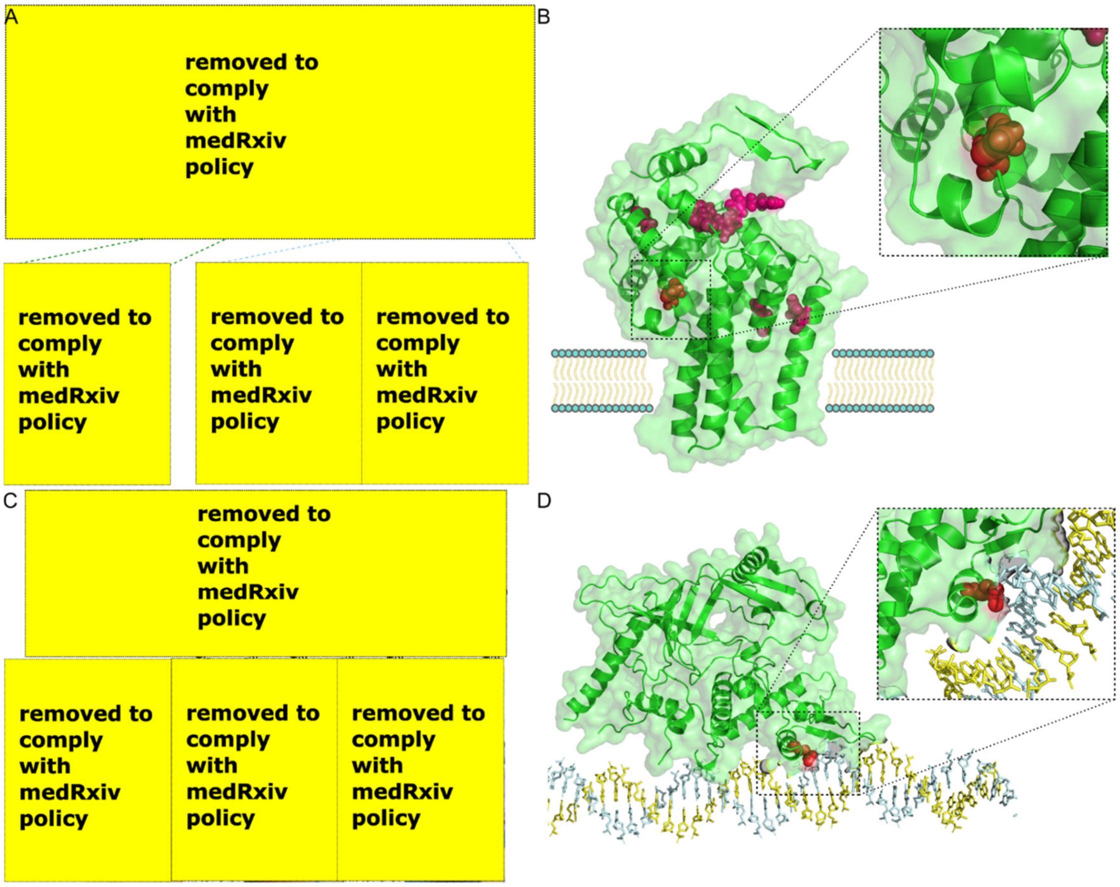


(**A**) Family MR049 with 3 branches ('a', 'b' and 'c'). In branch 'a' a variant in the candidate gene *CEP76* was identified (no facial photo available). In branch 'b' we identified a homozygous pathogenic variant in *DEGS1* in the affected girl (MR049-07, picture of affected individual with green border). A *de novo* pathogenic *ZEB2* variant was found in the affected individual in branch 'c' (MR049-13, blue border). He has typical facial features of Mowat-Wilson syndrome, with cupped ears, uplifted lobes, and central depression. (**B**) A protein homology model of *DEGS1* (generated using trRosetta) with the amino acid residue Asn255 highlighted in red. This residue is mutated by the missense variant c.764A>G, p.(Asn255Ser) identified in individual MR049-07 from the 'b' branch. Residues affected by other known pathogenic variants are colored in magenta. The endoplasmic reticulum (ER) membrane is schematically depicted. AA position 255 is highly conserved and located in the cytosolic domain of the sphingolipid desaturase DEGS1. The change of asparagine to a smaller serine residue likely disrupts the conformation of the enzymatic center characterized by histidine motifs [46]. (**C**) In Family MR073, the likely pathogenic variant c.2590C>T, p.(Arg864Trp) in *TAF1* was identified in 3 affected brothers in a hemizygous (+/y) state, while the mother and the unaffected siblings are heterozygous (+/-) or do not carry the variant (-/y), respectively. The affected brothers display a long philtrum, thin upper lip vermillion, microcephaly, and a wide nasal bridge. In the maternal family, no additional affected family members were reported. (**D**) The *TAF1* encoded protein TAFII250 (PDB 5FUR) interacts directly with the DNA. It is the largest subunit of the transcription factor IID complex (other subunits are not shown). The amino acid residue Arg864 (red) directly interacts with the DNA double helix and therefore seems essential. Please see the Supplementary methods for details of these analyses.

**Figure S2 | Clustering exomes into groups for CN analysis**

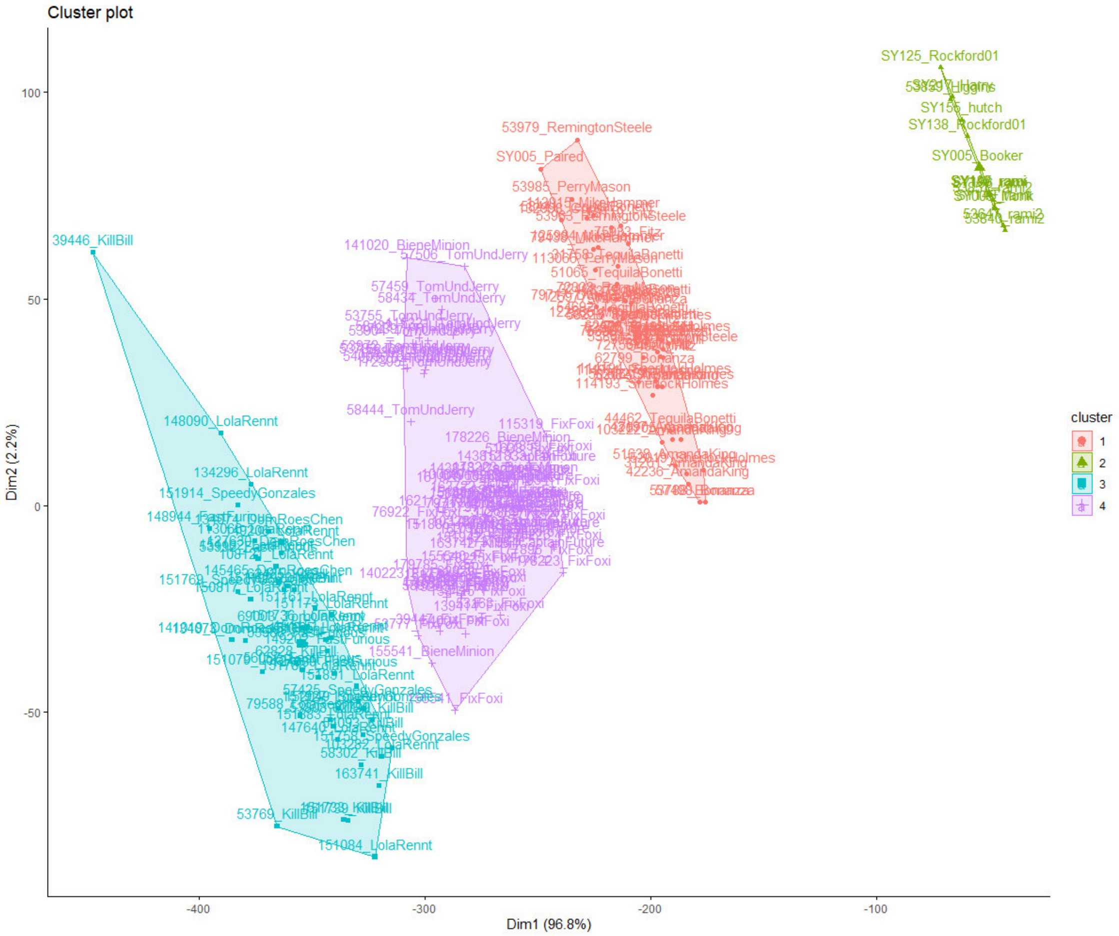


The plot ('fviz_cluster' command) of the k-means clustering using BAM coverage statistics indicates four clusters. The first principal component (Dim1, x-axis) explains most of the variation and likely corresponds to the combination of sequencing platform, sequencing read type (paired-end vs. single-end) and enrichment kits used. SOLiD exomes are in clusters 1 (single-end) and 2 (paired-end), while clusters 3 and 4 represent Illumina exomes with different enrichment kits. Note that clusters 2 and 3 could potentially be sub-clustered into two separate clusters each, but we decided against this option as we believe that more controls per cluster would, in this case, result in better results. The second principal component (Dim2, y-axis) is likely best correlated with the overall sequencing depth of the respective exome. Each dot represents a BAM file and is labeled with the sample's DNA identifier and original run name.

**Figure S3 | Ambiguous alignment in hg38 for *CBS***

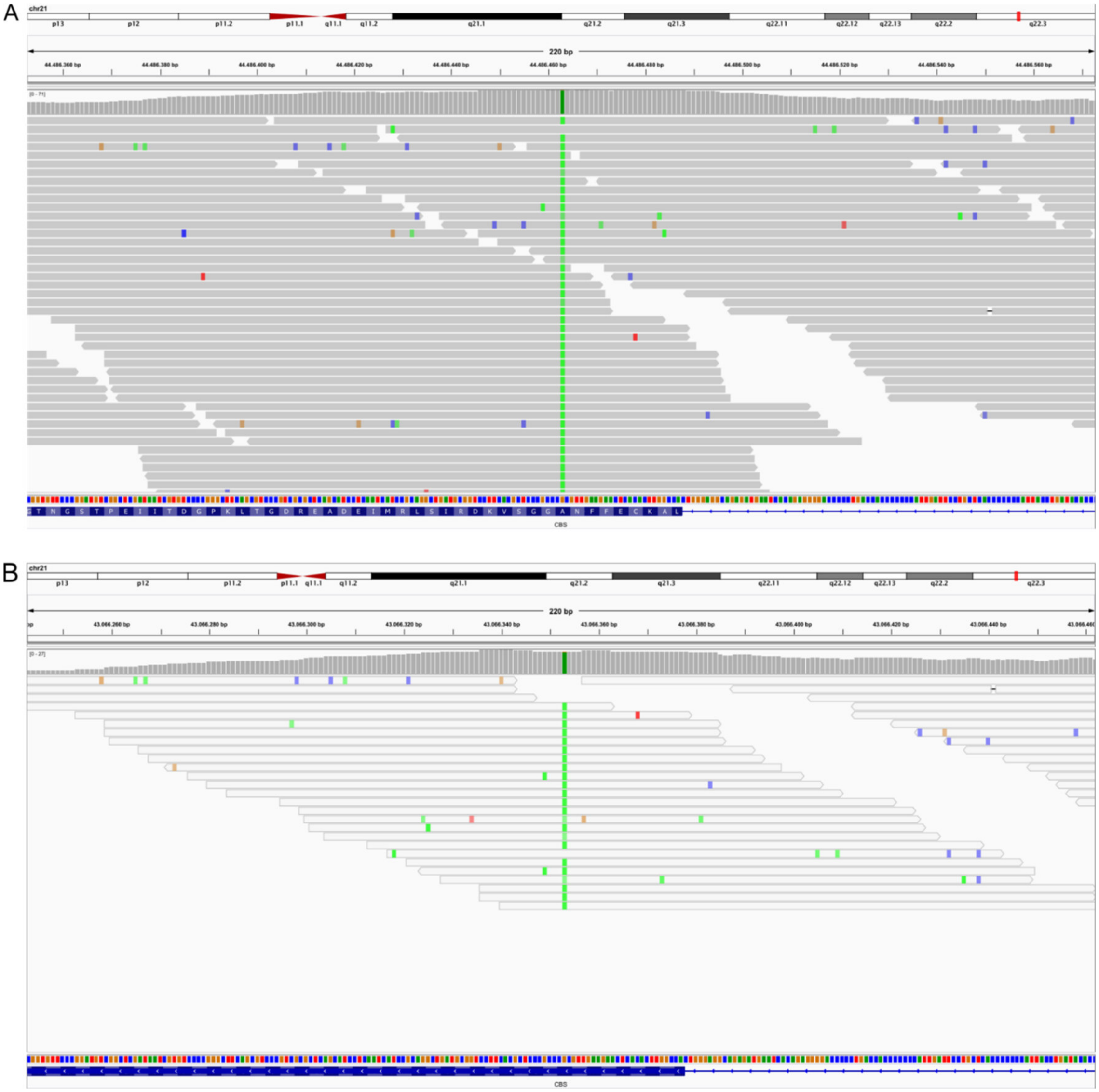


(**A**) IGV screenshots of the homozygous pathogenic *CBS* missense variant c.341C>T, p.(Ala114Val) which was previously called in the hg19 reference and reported. (**B**) It was not called from the realigned data due to a *CBS*/ *CBSL* duplication in the hg38 reference used, resulting in non-unique alignments of the reads.

**Figure S4 | Complex indel in *FAR1* missed in realigned data**

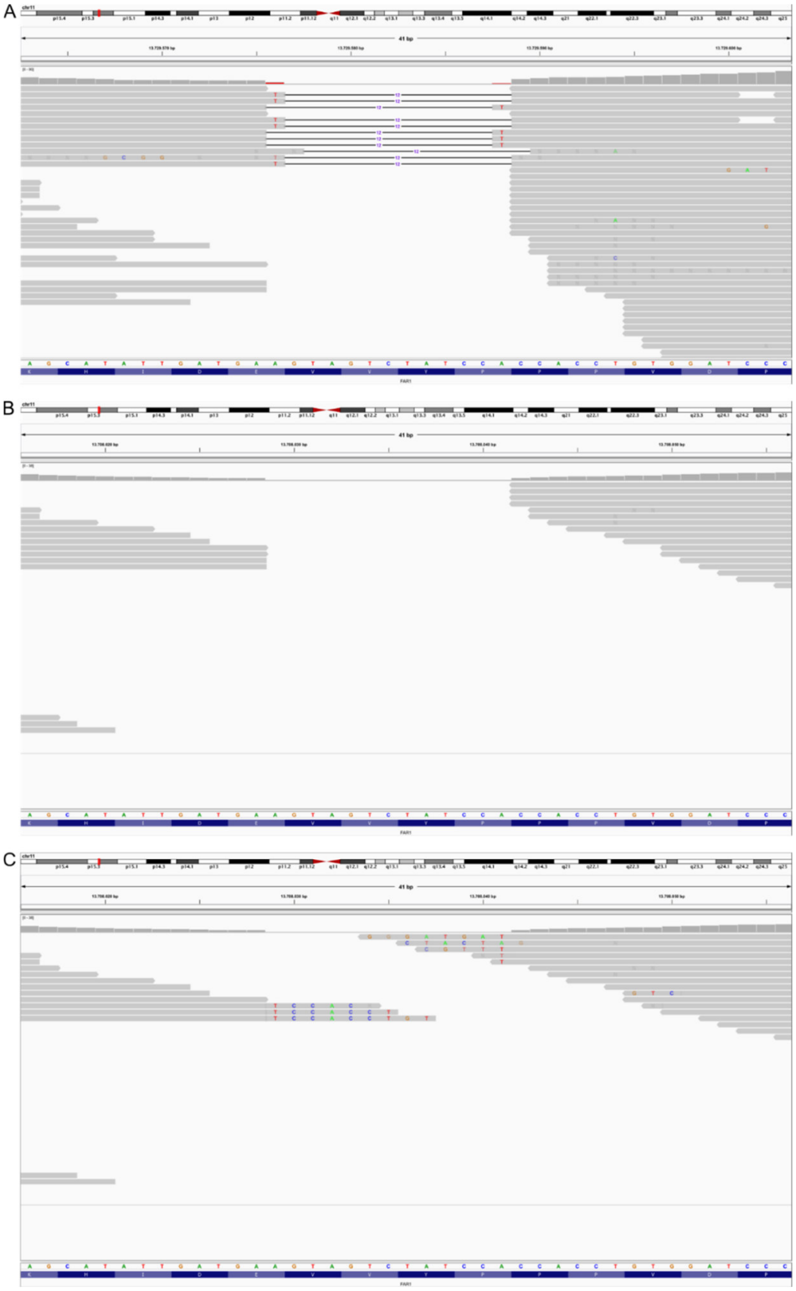

IGV screenshots of the homozygous *FAR1* in-frame indel variant c.495_507delinsT, p.(Glu165_Pro169delinsAsp) was previously called in the hg19 reference (**A**) and reported as pathogenic (see main text for discussion of classification for this gene and the variant after our re-evaluation). It was not called in the realignment because the new mapping tool (novoalignCS) could not correctly map the old color-space reads covering this variant (**B**). (**C**) shows the same region in the BAM file with soft-clipping information. The soft-clipped bases on the left side match the right deletion site and can be visually re-aligned to the reference sequence. The soft-clipped base information on the right hand cannot be visually re-aligned. Likely, the soft-clipped read part was too short for re-alignment through HaplotypeCaller.

**Figure S5 | Low coverage in exon 1 of C12orf57 hinders correct zygosity calling**

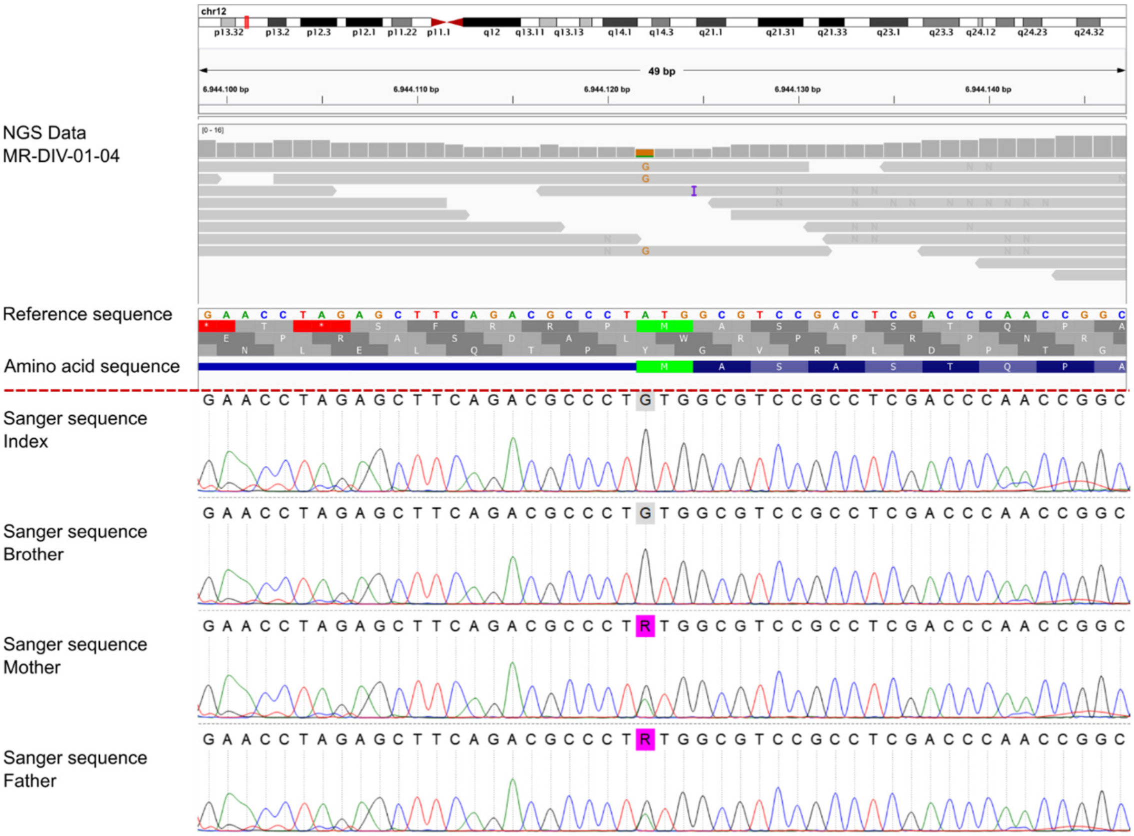


In Family MR-DIV-01 with two affected brothers with seizures, severe ID, muscular hypotonia, and short stature initially a variant in the candidate gene *FNDC3A* (NM_001079673.1:c.1186G>A, p.(Asp396Asn)) was reported. During reanalysis of this family, we identified the known pathogenic start loss variant in *C12orf57* (NM_001301834.1:c.1A>G, p.(Met1?)). The upper panel shows the sequencing data of the start codon in C12orf57 visualized in IGV. The first amino acid position is covered by four low quality reads. The zygosity of the identified variant could not be reliably determined. Sanger sequencing (lower panel) of the start codon in *C12orf57* revealed a homozygous state of the start loss variant c.1A>G, p.(Met1?) in the two affected brothers (top two Sanger traces). Both parents are heterozygous carriers (bottom two Sanger traces).

**Figure S6 | Segregation of the C-terminal frameshifting *ASXL3* variant in pedigree MR-SYR-14**

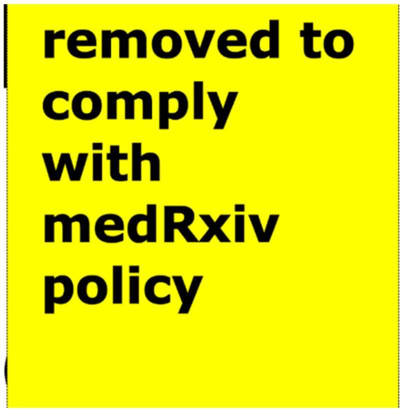


In family MR-SYR-14, two affected siblings with mild ID, microcephaly, aggressive behavior, and self-mutilation the frameshift deletion c.4462_4465delACTG, p.(Thr1488Serfs*17) was identified (+) in *ASXL3* (NM_030632.2) in both siblings. Interpretation of this variant was inconclusive since truncating variants further downstream were reported to be pathogenic (e.g. p.Val1505Aspfs*3)[34] and thus causative for Bainbridge-Ropers syndrome (#615485). On the other hand, the unaffected father in the MR-SYR-14 family carries the variant in an apparent heterozygous state in DNA derived from peripheral blood. A postzygotic mosaic in the father, as described for *ASXL3* [34(p3)] , could explain this observation.

### SUPPLEMENTARY TABLES

#### Table S1 | Primer sequences for segregation analysis

| Primer name | sequence | Family ID |
|---|---|---|
| TAF1_MR073_F | CACATCGGGAAAGATGAAAATAC | MR073 |
| TAF1_MR073_R | GATCTGCTACCTCCTTTCAACAG | MR073 |
| ADNP_MR333_F | AAGCATCCTCAGGAATTACCTTC | MR333 |
| ADNP_MR333_R | TAGCAGCCAGTTTATGGTTATGG | MR333 |
| GRIN2A_MR136_F | ATGCCGAGAGTCAATTTCTGTAA | MR136 |
| GRIN2A_MR136_R | ATGCATTTACCTCCTAACACCAG | MR136 |
| ZNF143_MR071a_F | ATTTCCAACACCATTACATCCTC | MR071a |
| ZNF143_MR071a_R | TGCTAAATCAGGAACTAGGAAGTTG | MR071a |
| ASXL3_MR014_F | AAAATTCAGGGCCTCGAAAC | MR-SYR-14 |
| ASXL3_MR014_R | ATACCTTCGTTGGCAACTGG | MR-SYR-14 |
| HNRNPH2_MR-TUR-05_F | TACAGGTCCGAATAGCCCTG | MR-TUR-05 |
| HNRNPH2_MR-TUR-05_R | TCTGCCTCACCGGTAACTCT | MR-TUR-05 |
| C12orf57 | TGTAGGACGTGGCTCTTTAT | MR-DIV-01 |
| C12orf57 | TTATCCCGAGCCTCGTCCAT | MR-DIV-01 |
| ZEB2_MR049c_F | TTGGTAAAATGGGAAGGTTTTG | MR049c |
| ZEB2_MR049c_R | CAGGAATTTGTGAAGGAATGG | MR049c |

### SUPPLEMENTARY DATA FILES

These data files used for analyses and tables/ figures are available for download from Zenodo:

**File S2**: comprehensive tabular data describing relevant cohort information, including relevant variants, families, individuals, sequenced samples, and analyzed BAM files [35]

**File S3**: comprehensive variant data including all filtered and reviewed homozygous, dominant, recessive variant, copy number calls and runs of homozygosity [35]

### REFERENCES


1. Tange O. *Gnu Parallel 2018*. Zenodo; 2018. doi:10.5281/ZENODO.1146014
2. Li H, Handsaker B, Wysoker A, et al. The Sequence Alignment/Map format and SAMtools. *Bioinformatics*. 2009;25(16):2078-2079. doi:10.1093/bioinformatics/btp352
3. McKenna A, Hanna M, Banks E, et al. The Genome Analysis Toolkit: A MapReduce framework for analyzing next-generation DNA sequencing data. *Genome Res*. 2010;20(9):1297-1303. doi:10.1101/gr.107524.110
4. DePristo MA, Banks E, Poplin R, et al. A framework for variation discovery and genotyping using next-generation DNA sequencing data. *Nat Genet*. 2011;43(5):491-498. doi:10.1038/ng.806
5. Li H. Aligning sequence reads, clone sequences and assembly contigs with BWA-MEM. *ArXiv13033997 Q-Bio*. Published online May 26, 2013. Accessed November 30, 2020. http://arxiv.org/abs/1303.3997
6. Zhao H, Sun Z, Wang J, Huang H, Kocher JP, Wang L. CrossMap: a versatile tool for coordinate conversion between genome assemblies. *Bioinformatics*. 2014;30(7):1006-1007. doi:10.1093/bioinformatics/btt730
7. Okonechnikov K, Conesa A, García-Alcalde F. Qualimap 2: advanced multi-sample quality control for high-throughput sequencing data. *Bioinformatics*. Published online October 1, 2015:btv566. doi:10.1093/bioinformatics/btv566
8. Poplin R, Ruano-Rubio V, DePristo MA, et al. *Scaling Accurate Genetic Variant Discovery to Tens of Thousands of Samples*. Genomics; 2017. doi:10.1101/201178
9. Deutsch P. *GZIP File Format Specification Version 4.3*. RFC Editor; 1996:RFC1952. doi:10.17487/rfc1952
10. Li H. Tabix: fast retrieval of sequence features from generic TAB-delimited files. *Bioinformatics*. 2011;27(5):718-719. doi:10.1093/bioinformatics/btq671
11. Cingolani P, Platts A, Wang LL, et al. A program for annotating and predicting the effects of single nucleotide polymorphisms, SnpEff: SNPs in the genome of Drosophila melanogaster strain w1118; iso-2; iso-3. *Fly (Austin)*. 2012;6(2):80-92. doi:10.4161/fly.19695
12. Cingolani P, Patel VM, Coon M, et al. Using Drosophila melanogaster as a Model for Genotoxic Chemical Mutational Studies with a New Program, SnpSift. *Front Genet*. 2012;3. doi:10.3389/fgene.2012.00035
13. Liu X, Jian X, Boerwinkle E. dbNSFP: a lightweight database of human nonsynonymous SNPs and their functional predictions. *Hum Mutat*. 2011;32(8):894-899. doi:10.1002/humu.21517
14. Xiong HY, Alipanahi B, Lee LJ, et al. The human splicing code reveals new insights into the genetic determinants of disease. *Science*. 2015;347(6218):1254806-1254806. doi:10.1126/science.1254806
15. Jian X, Boerwinkle E, Liu X. In silico prediction of splice-altering single nucleotide variants in the human genome. *Nucleic Acids Res*. 2014;42(22):13534-13544. doi:10.1093/nar/gku1206
16. Rentzsch P, Witten D, Cooper GM, Shendure J, Kircher M. CADD: predicting the deleteriousness of variants throughout the human genome. *Nucleic Acids Res*. 2019;47(D1):D886-D894. doi:10.1093/nar/gky1016
17. Karczewski KJ, Francioli LC, Tiao G, et al. The mutational constraint spectrum quantified from variation in 141,456 humans. *Nature*. 2020;581(7809):434-443. doi:10.1038/s41586-020-2308-7
18. Taliun D, Harris DN, Kessler MD, et al. *Sequencing of 53,831 Diverse Genomes from the NHLBI TOPMed Program*. Genomics; 2019. doi:10.1101/563866
19. Landrum MJ, Lee JM, Riley GR, et al. ClinVar: public archive of relationships among sequence variation and human phenotype. *Nucleic Acids Res*. 2014;42(Database issue):D980-985. doi:10.1093/nar/gkt1113
20. Abramovs N, Brass A, Tassabehji M. GeVIR is a continuous gene-level metric that uses variant distribution patterns to prioritize disease candidate genes. *Nat Genet*. 2020;52(1):35-39. doi:10.1038/s41588-019-0560-2

21. Kochinke K, Zweier C, Nijhof B, et al. Systematic Phenomics Analysis Deconvolutes Genes Mutated in Intellectual Disability into Biologically Coherent Modules. *Am J Hum Genet*. 2016;98(1):149-164. doi:10.1016/j.ajhg.2015.11.024
22. Robinson JT, Thorvaldsdóttir H, Winckler W, et al. Integrative genomics viewer. *Nat Biotechnol*. 2011;29(1):24-26. doi:10.1038/nbt.1754
23. Hamosh A, Scott AF, Amberger JS, Bocchini CA, McKusick VA. Online Mendelian Inheritance in Man (OMIM), a knowledgebase of human genes and genetic disorders. *Nucleic Acids Res*. 2005;33(Database issue):D514-517. doi:10.1093/nar/gki033
24. Talevich E, Shain AH, Botton T, Bastian BC. CNVkit: Genome-Wide Copy Number Detection and Visualization from Targeted DNA Sequencing. *PLOS Comput Biol*. 2016;12(4):e1004873. doi:10.1371/journal.pcbi.1004873
25. Firth HV, Richards SM, Bevan AP, et al. DECIPHER: Database of Chromosomal Imbalance and Phenotype in Humans Using Ensembl Resources. *Am J Hum Genet*. 2009;84(4):524-533. doi:10.1016/j.ajhg.2009.03.010
26. Li H. A statistical framework for SNP calling, mutation discovery, association mapping and population genetical parameter estimation from sequencing data. *Bioinformatics*. 2011;27(21):2987-2993. doi:10.1093/bioinformatics/btr509
27. Untergasser A, Cutcutache I, Koressaar T, et al. Primer3--new capabilities and interfaces. *Nucleic Acids Res*. 2012;40(15):e115. doi:10.1093/nar/gks596
28. Okonechnikov K, Golosova O, Fursov M. Unipro UGENE: a unified bioinformatics toolkit. *Bioinformatics*. 2012;28(8):1166-1167. doi:10.1093/bioinformatics/bts091
29. Louder RK, He Y, López-Blanco JR, Fang J, Chacón P, Nogales E. Structure of promoter-bound TFIID and model of human pre-initiation complex assembly. *Nature*. 2016;531(7596):604-609. doi:10.1038/nature17394
30. Yang J, Anishchenko I, Park H, Peng Z, Ovchinnikov S, Baker D. Improved protein structure prediction using predicted interresidue orientations. *Proc Natl Acad Sci*. 2020;117(3):1496-1503. doi:10.1073/pnas.1914677117
31. Mirzaa GM, Chong JX, Piton A, et al. De novo and inherited variants in ZNF292 underlie a neurodevelopmental disorder with features of autism spectrum disorder. *Genet Med Off J Am Coll Med Genet*. 2020;22(3):538-546. doi:10.1038/s41436-019-0693-9
32. Lin Y, Zeng C, Lu Z, Lin R, Liu L. A novel homozygous splice-site variant of NCAPD2 gene identified in two siblings with primary microcephaly: The second case report. *Clin Genet*. 2019;96(1):98-101. doi:10.1111/cge.13559
33. Martin CA, Murray JE, Carroll P, et al. Mutations in genes encoding condensin complex proteins cause microcephaly through decatenation failure at mitosis. *Genes Dev*. 2016;30(19):2158-2172. doi:10.1101/gad.286351.116
34. Schirwani S, Hauser N, Platt A, et al. Mosaicism in ASXL3-related syndrome: Description of five patients from three families. *Eur J Med Genet*. 2020;63(6):103925. doi:10.1016/j.ejmg.2020.103925
35. Popp, Bernt, Bartolomaeus, Tobias. Data files for manuscript “Re-evaluation and Re-analysis of 152 research exomes five years after the initial report reveals clinically relevant changes in 20%.” Published online September 23, 2022. doi:10.5281/zenodo.7113408
